# Comprehensive Analysis of Metabolic Reprogramming-Associated Key Genes and Immune Microenvironment in Heart Failure with Preserved Ejection Fraction

**DOI:** 10.1101/2025.01.15.25320619

**Authors:** Na Yan, Jiangyong Yang

## Abstract

**Background:** Heart failure with preserved ejection fraction (HFpEF) represents a profoundly heterogeneous cardiovascular condition; characterized by intricate molecular pathways that have not yet been fully elucidated. This study seeks to uncover metabolic reprogramming-associated key genes (Key Genes) linked to HFpEF, investigate their functional significance and influence on the immune microenvironment, and establish a robust early diagnostic framework.

**Methods:** We obtained datasets GSE194151 and GSE180065 from the GEO database. In order to counteract batch effects, metabolic reprogramming-related differentially expressed genes (MRRDEGs) were identified by differential expression analysis using the sva R package. Enrichment analysis using Gene Ontology (GO) and the Kyoto Encyclopedia of Genes and Genomics (KEGG) elucidated the associated biological functions and pathways. Gene set enrichment analysis (GSEA) and gene set variation analysis (GSVA) were used to examine biological differences within subgroups using risk assessment. Single sample gene set enrichment analysis (ssGSEA) was used to measure immune cell infiltration. Protein-protein interaction networks (PPI Networks), mRNA-TF, mRNA-RBP, and mRNA-miRNA interaction networks were used to investigate multilevel regulatory mechanisms. A diagnostic model for HFpEF, constructed using logistic regression, SVM, and LASSO, was validated through ROC analysis and external datasets.

**Results:** In total, 34 MRRDEGs were identified, significantly enriched in processes like fatty acid metabolism and circadian rhythm regulation. A diagnostic model comprising five Key Genes(*Hnrnpd*, *Hpcal1*, *Hadha*, *Hspd1*, and *Naa10*) showed strong performance, with *Hspd1* achieving an Area Under Curve (AUC) > 0.9. The validation using GSE180065 and HFpEF_2020 confirmed the reliability of the model. Immune analysis revealed significant enrichment of Activated CD4 T cells, type 2 T helper cells, and macrophages in the high-risk groups, correlating with specific Key Genes (p < 0.05). GSEA and GSVA linked high-risk groups to TGF-β signaling and fatty acid oxidation. PPI and regulatory network analyses further underscored the functional importance of Key Genes.

**Conclusion:** Our study systematically identified metabolic reprogramming-related Key Genes and their associations with immune microenvironment changes in HFpEF. These findings reveal the potential mechanisms underlying HFpEF and offer a high-performance diagnostic model, providing novel insights and therapeutic targets for molecular studies and immunotherapy in HFpEF.

## 1 Introduction

Heart failure (HF) with preserved ejection fraction (HFpEF) has emerged as a significant clinical and public health concern, particularly as its prevalence continues to rise in the aging population(1). This condition, characterized by signs and symptoms of HF alongside a left ventricular ejection fraction (LVEF) of 50% or higher, presents unique challenges in both diagnosis and management. The complex interplay of comorbidities, such as hypertension, diabetes, and obesity, further complicates the clinical landscape, leading to substantial morbidity and mortality associated with HFpEF (2). Despite increasing recognition, the underlying pathophysiology of HFpEF remains poorly understood, primarily because of its heterogeneous nature and multifactorial contributions to its development (3) (4).Current therapeutic options for HFpEF are limited, with no pharmacological treatment demonstrating significant efficacy in improving outcomes (5, 6). Existing management strategies largely focus on symptom relief and treatment of associated comorbidities; however, they fail to address the fundamental mechanisms driving the disease(7). Therefore, it is crucial to develop novel diagnostic methods and therapeutic strategies that target the distinctive characteristics of HFpEF(8).

Recent findings have drawn attention to metabolic reprogramming and alterations in cellular energy metabolism as critical factors in HFpEF progression(9). Nevertheless, the specific genes and pathways involved; as well as their contributions to disease pathology; are yet to be fully understood (10).The interplay between metabolic reprogramming and the immune microenvironment has emerged as a crucial area of research in HFpEF(11). Aberrant immune cell infiltration, which is known to mediate inflammation and cardiac remodeling, has been implicated in the pathogenesis of the disease (12, 13). This highlights the need to investigate the interactions between metabolic and immune alterations in HFpEF(14). Despite growing evidence, the absence of reliable biomarkers and predictive models for early HFpEF detection presents a major challenge, underscoring the urgent need for integrative approaches that incorporating molecular and immune profiling.

This study focused on identifying the metabolism-reprogramming-related genes (MRRDEGs) implicated in HFpEF and examining their regulatory roles in the immune microenvironment. By leveraging comprehensive bioinformatics analyses, including differential expression profiling, functional enrichment, and immune infiltration assessments, this study aimed to elucidate the molecular underpinnings of HFpEF. In addition, the development and verification of a diagnostic framework based on essential metabolic genes provides important perspectives for the early and accurate detection of HFpEF, which could have significant implications for clinical applications.

## 2 Materials and Methods

### 2.1 Data Collection and Preprocessing

The HFpEF datasets GSE194151 and GSE180065 were retrieved from the GEO database (https://www.ncbi.nlm.nih.gov/geo/) using the GEOquery R package (15). GSE194151 contains cardiac tissue sequencing data from 15 HFpEF samples induced by a high-fat diet and 15 control samples from a normal diet. GSE180065 includes cardiac tissue sequencing data from 10 HFpEF samples and 5 control samples. All samples were derived from mice (Mus musculus); and the platform used was GPL24247 (see Table 1). The data format is in Counts, and the data were standardized using Fragments Per Kilobase Per Million (FPKM) format for subsequent analysis. External validation dataset HFpEF_2020, sourced from Hahn et al. (16), includes 41 HFpEF samples, 59 samples of heart failure with reduced ejection fraction (HFrEF), and 45 control samples. In this analysis, all HFrEF samples were excluded, and only HFpEF and control samples were included.

**Table 1.** GEO Microarray Chip Information.

|  | GSE194151 | GSE180065 |
| --- | --- | --- |
| Platform | GPL24247 | GPL24247 |
| Species | Mus musculus | Mus musculus |
| Tissue | Heart Tissue | Heart Tissue |
| Samples in HFpEF group | 15 | 10 |
| Samples in Control group | 15 | 5 |
| Reference | PMID:35787630 | / |

GEO, Gene Expression Omnibus; HFpEF, Heart Failure with Preserved Ejection Fraction.

A list of Metabolic Reprogramming-related Genes (MRRGs) was collected from the GeneCards database (17); using the keyword "Metabolic Reprogramming" and limited to protein-coding genes, resulting in 1,423 MRRGs. Human-mouse gene ID conversion was performed using the *homologene* R package, which retained 1,327 MRRGs. Additionally, gene lists from the PubMed database (18)were incorporated, resulting in a final total of 1,328 unique MRRGs. To eliminate batch effects, the R package *sva* (19) was employed to perform batch correction on the datasets GSE194151 and GSE180065. Data standardization and normalization were performed using the R package *limma* (20). The workflow of this study is presented in Figure 1.

**Fig. 1.**
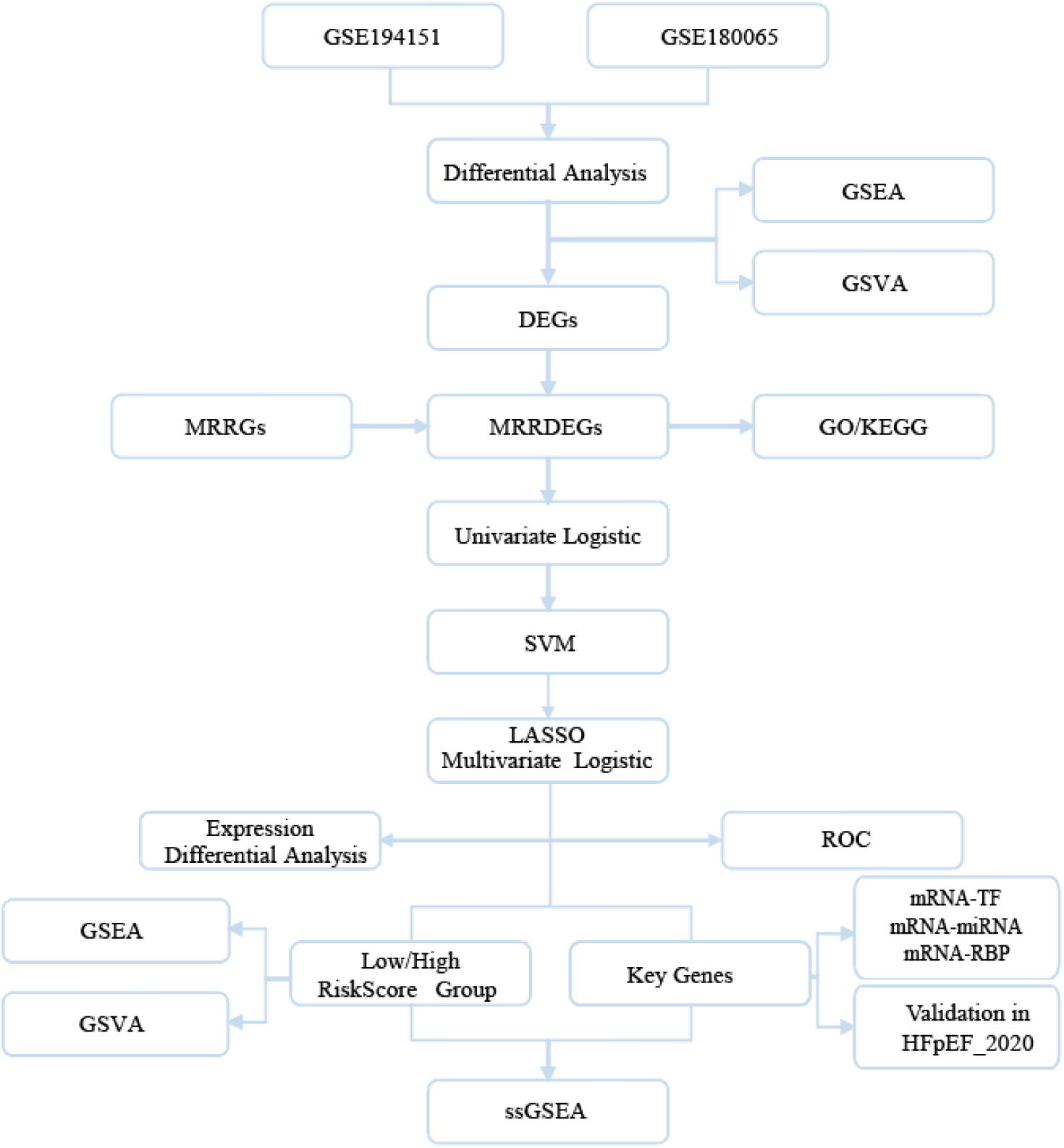
Technology Roadmap.

### 2.2 Differential Expression Gene (DEG) Analysis

The samples were categorized into HFpEF and control groups according to the grouping information from GSE194151 and GSE180065. DEG analysis was performed using the R package *limma* with thresholds set at |logFC| > 0 and p-value < 0.05. Genes with logFC > 0 and p-value < 0.05 were considered upregulated, while genes with logFC < 0 and p-value < 0.05 were considered downregulated. A volcano plot was generated using the *ggplot2* R package to visualize the differential expression results. To identify MRRDEGs, DEGs were intersected with MRRGs; and the results were displayed using a Venn diagram. A heatmap of the MRRDEGs expression was created using the *pheatmap* package.

### 2.3 Gene Ontology (GO) and Kyoto Encyclopedia of Genes and Genomes(KEGG) Enrichment Analysis

The *clusterProfiler* package was utilized to perform GO and KEGG pathway enrichment analyses for the MRRDEGs (21). The GO analysis included three distinct categories: biological process (BP), cellular component (CC), and molecular function (MF). The screening criteria were set to p-value < 0.05 and FDR < 0.25. KEGG analysis was used to explore the main pathways involved in MRRDEGs using the same screening criteria.

### 2.4 Gene Set Enrichment Analysis (GSEA)

GSEA was executed using the *clusterProfiler* R package to rank gene sets in the GSE194151 dataset, with significantly enriched gene sets selected based on an adjusted p-value < 0.05 and FDR < 0.25. Gene sets were sourced from the Molecular Signatures Database (MSigDB) version m2.all.v2023.2.Mm.symbols. The analysis parameters included a starting value of 2020 and 1,000 permutations.

### 2.5 Gene Set Variation Analysis (GSVA)

GSVA is an unsupervised method employed to examine variations in gene sets. It was applied to evaluate the differential expression of these gene sets among various samples. The MSigDB(22) gene set *c2.cp.v2023.2.Hs.symbols.gmt* was selected, and GSVA was used to compute functional enrichment disparities between the HFpEF and control groups, and between the high- and low-risk groups. The screening criterion was adjusted to a p-value < 0.05.

### 2.6 Construction of the HFpEF Diagnostic Model

Logistic regression was used to analyze the MRRDEGs, with genes having a p-value < 0.05 included in the diagnostic model. An Support Vector Machine (SVM ) model was constructed based on the selected MRRDEGs, and the model was optimized using Least Absolute Shrinkage and Selection Operator (LASSO) regression. The risk score (Risk Score) of the final model was calculated using the following formula:

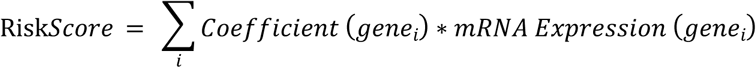

### 2.7 Model Validation

A nomogram was developed from the logistic regression model outcomes, and a calibration curve was created to evaluate the precision of the model. The clinical utility of the model was evaluated through Decision Curve Analysis (DCA) (23), and its diagnostic ability was assessed using the ROC curve and calculation of the Area Under Curve(AUC).

### 2.8 Validation of Key Genes Expression Differences

A comparative plot was created to illustrate the expression of Key Genes, and their diagnostic effectiveness was assessed by calculating the ROC curve and AUC.

### 2.9 Single-sample Gene Set Enrichment Analysis (ssGSEA) Immune Infiltration Analysis

Single-sample Gene Set Enrichment Analysis (ssGSEA) [24] was used to assess the relative abundance of immune cell populations across various groups, and a bubble plot was generated to illustrate the relationship between Key Genes and immune cell infiltration.

### 2.10 A protein-protein interaction (PPI) Network of Key Genes

A protein-protein interaction (PPI) network of Key Genes was developed utilizing the STRING database(24); and functional similarity was calculated using the R package GOSemSim(25). Similar genes were predicted using GeneMANIA(26).

### 2.11 mRNA-TF, mRNA-RBP, and mRNA-miRNA Interaction Networks

mRNA-TF (Transcription Factor), mRNA-RBP (RNA-Binding Protein), and mRNA-miRNA (MicroRNA) regulatory networks were constructed based on data from the ChIPBase(27) and StarBase(28) databases, and these networks were visualized using Cytoscape(29).

### 2.12 Statistical Analysis

Our study used R software (version 4.2.2) for data processing and statistical analysis. Unless otherwise stated, independent Student’s t tests were used to determine the significance of normally distributed variables for comparing continuous variables between two groups. For non-normally distributed variables, the Mann-Whitney U test (Wilcoxon rank-sum test) was applied to evaluate differences. Comparisons involving three or more groups were made using the Kruskal-Wallis test. Spearman correlation analysis was used to calculate the correlation coefficients between different molecules. All p-values were two-sided unless stated otherwise, with a p-value of less than 0.05 considered indicative of statistical significance.

## 3 Results

### 3.1 Batch Effect Correction for HFpEF Datasets

To improve data reliability, we first applied batch effect removal to the HFpEF datasets GSE194151 and GSE180065 using the *sva* R package. Comparison of boxplots before and after batch effect removal (GSE194151: Fig. 2A-B; GSE180065: Fig. 2C-D) demonstrated a significant improvement in the consistency of sample distribution between the two datasets after correction, providing a solid data foundation for subsequent analyses.

**Fig. 2.**
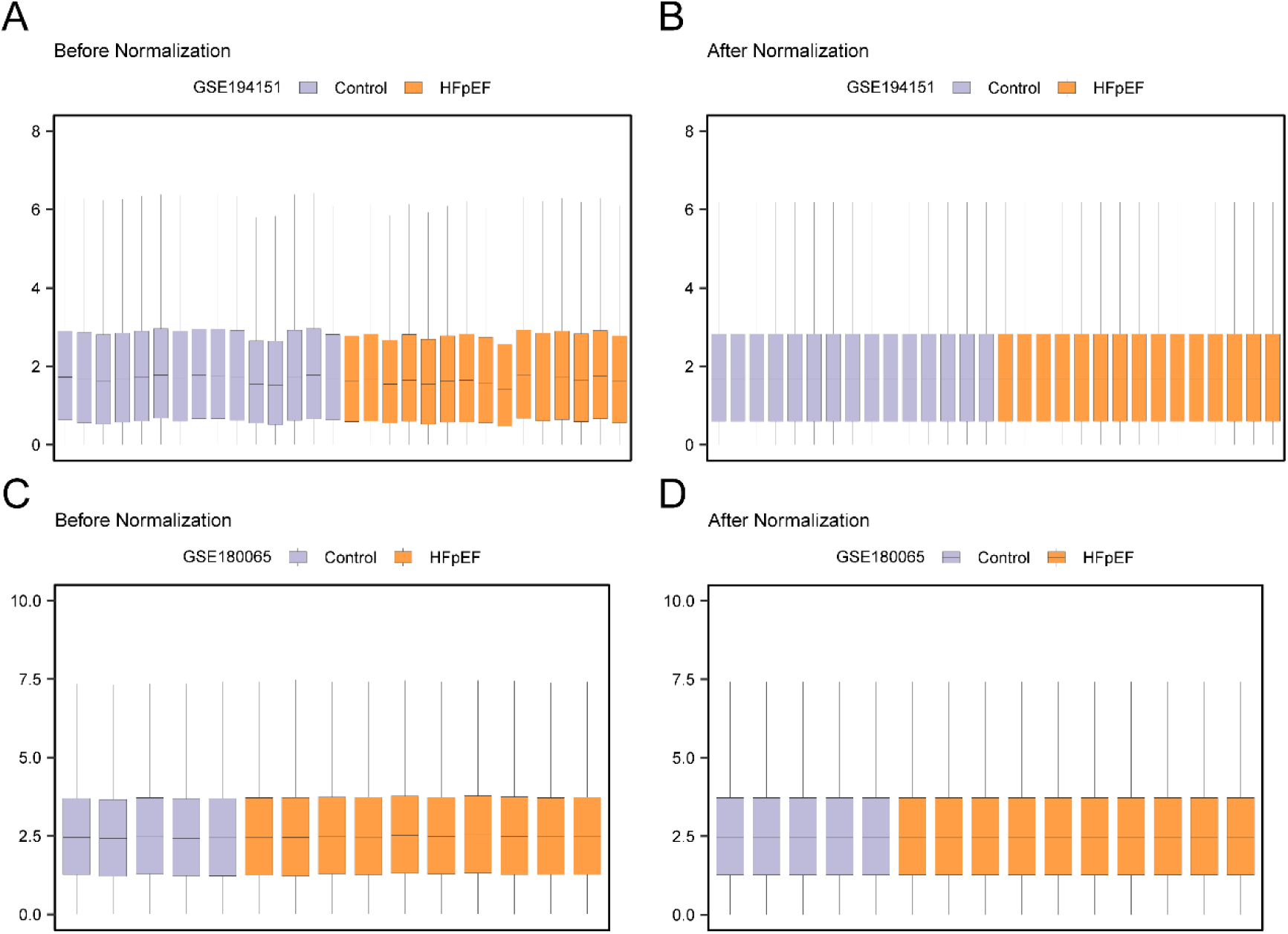
Debatching of the data. A. Box plot of GSE194151 distribution of dataset before going batch. B. The boxplot of GSE194151 distribution of the dataset after going to batch processing. C. Boxplot of GSE180065 distribution of dataset before batch processing. D. The boxplot of GSE180065 distribution of the dataset after batch processing. HFpEF, Heart Failure with Preserved Ejection Fraction. Orange is the HFpEF group, light blue is the Control group.

### 3.2 Identification of Metabolic Reprogramming-Related Differentially Expressed Genes in HFpEF

DEG analysis was conducted between the HFpEF and control groups in the GSE194151 and GSE180065 datasets using the *limma* R package. In the GSE194151 dataset, 1,236 DEGs were detected, with 541 upregulated and 695 downregulated genes (Fig. 3A). In the GSE180065 dataset, 2,469 DEGs were identified, including 1,135 upregulated and 1,334 downregulated genes (Fig. 3B). To further identify MRRDEGs, we intersected the DEGs from both datasets with a known set of metabolic reprogramming-related genes, identifying 34 MRRDEGs (e.g., *Pparg*, *Mthfd2*, *Hadha*), which are displayed in a Venn diagram (Fig. 3C).

**Fig. 3.**
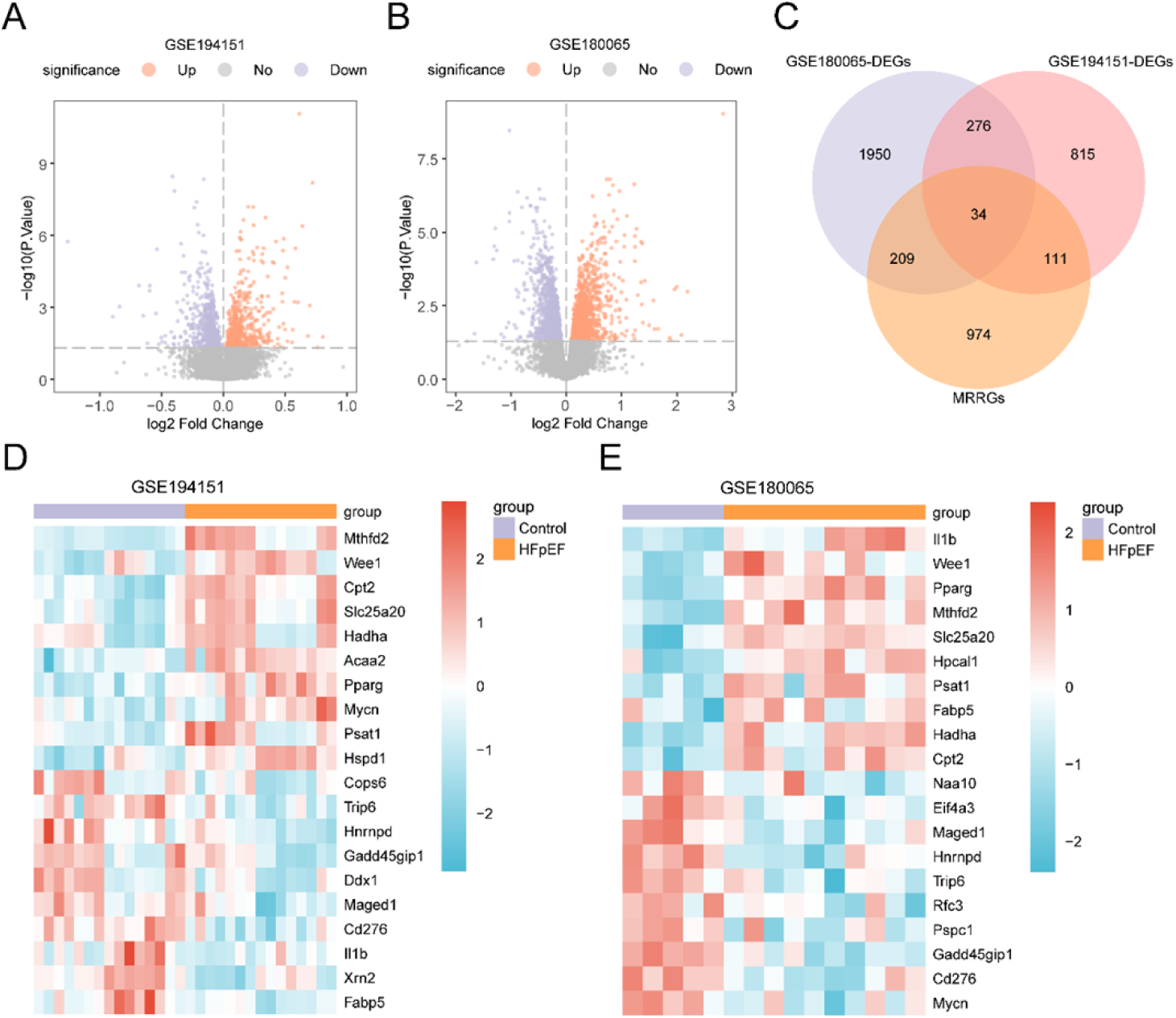
Differential gene expression analysis. A-B. Volcano plot of differentially expressed genes analysis between heart failure with preserved ejection fraction (HFpEF) group and Control (Control) group in datasets GSE194151 (A) and GSE180065 (B). C. Venn diagram of genes and metabolic reprogramming-related genes (MRRGs) in all heart failure samples with preserved ejection fraction from differentially expressed genes (DEGs) in datasets GSE194151 and GSE180065. D-E. Heat map of metabolic reprogramme-associated differentially expressed genes (MRRDEGs) in datasets GSE194151 (D) and GSE180065 (E). HFpEF, Heart Failure with Preserved Ejection Fraction; DEGs, Differentially Expressed Genes; MRRGs, Metabolic Reprogramming Related Genes; MRRDEGs, Metabolic Reprogramming-Related Differentially Expressed Genes. Orange is the heart failure with preserved ejection fraction (HFpEF) group, and light blue is the Control group. In the heat map, red represents high expression and blue represents low expression.

We further analyzed the expression patterns of the MRRDEGs by selecting the top 10 upregulated and downregulated genes with the highest and lowest logFC values, respectively. A heatmap was generated to visualize the expression differences between groups (Fig. 3D-E), highlighting the specific expression of these genes in HFpEF, and further elucidating their potential functional roles.

### 3.3 GO and KEGG Enrichment Analyses Reveal HFpEF-Related Metabolic Pathways

To gain further insights into the biological functions of MRRDEGs, we performed GO and KEGG enrichment analyses. The findings revealed that these genes are significantly associated with biological processes (BP) such as circadian rhythm regulation and fatty acid oxidation. Cellular component (CC) analysis showed their localization in the mitochondrial matrix and U2-type spliceosomes, while molecular function (MF) analysis revealed associations with ribonucleoprotein complex binding and acyltransferase activity. KEGG pathway analysis further highlighted notable enrichment in pathways associated to fatty acid metabolism, the PPAR signaling pathway, and branched-chain amino acid degradation (Table 2). These findings are represented visually in a bubble chart (Fig. 4A), and a network diagram (Fig. 4B-E), suggesting that metabolic reprogramming might play an important role in the pathophysiology of HFpEF.

**Fig. 4.**
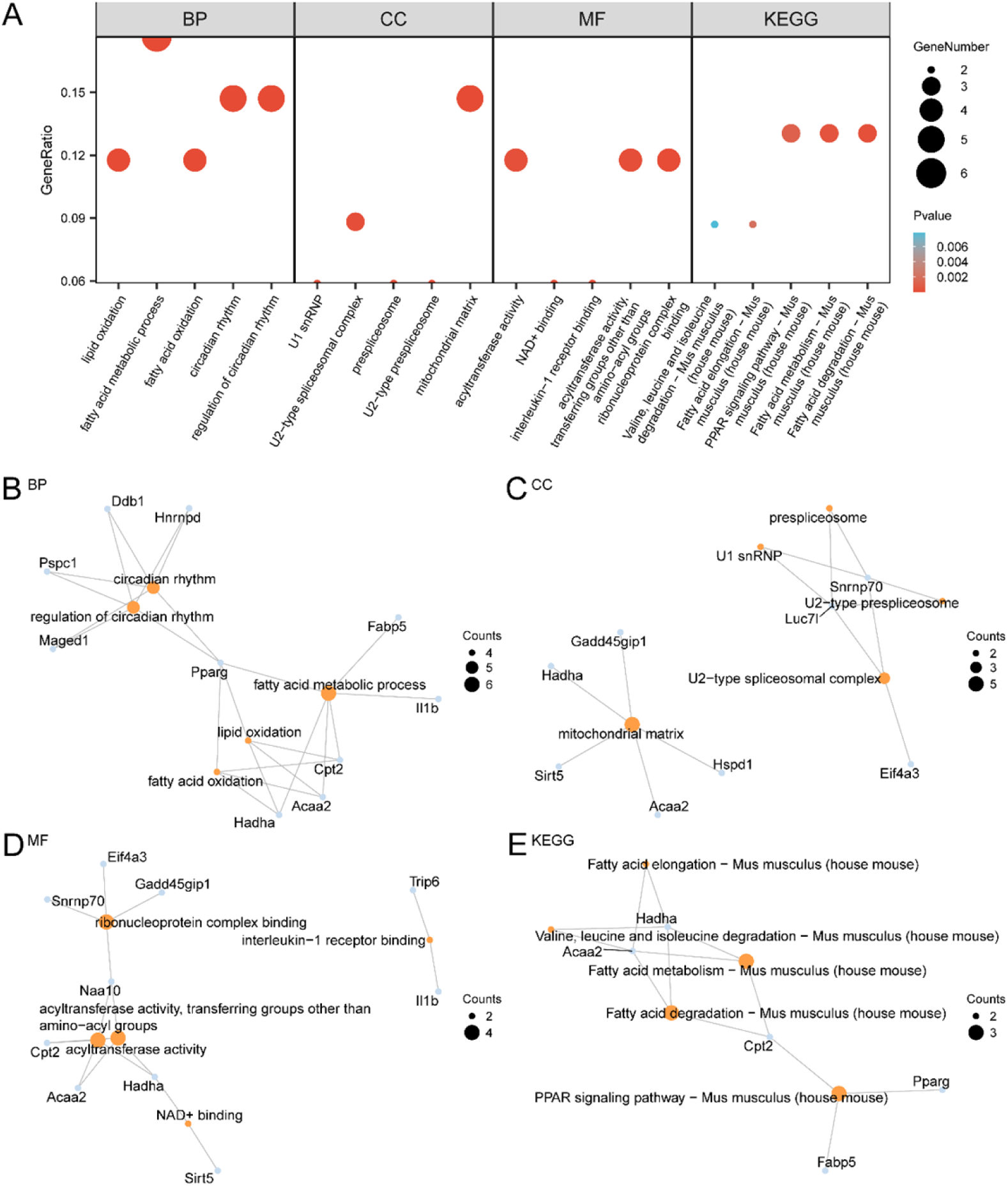
GO and KEGG enrichment analysis of MRRDEGs. A.Bubble plot of gene ontology (GO) and pathway (KEGG) enrichment analysis results of metabolic reprogramming related differentially expressed genes (MRRDEGs) : biological process (BP), cellular component (CC), molecular function (MF) and biological pathway (KEGG). GO terms and KEGG terms are shown on the abscissa. B-E. Gene ontology (GO) and pathway (KEGG) enrichment analysis results of MRRDEGs network diagram showing: BP (B), CC (C), MF (D) and KEGG (E). The orange nodes represent items, the blue nodes represent molecules, and the lines represent the relationship between items and molecules. MRRDEGs, Metabolic Reprogramming-Related Differentially Expressed Genes; GO, Gene Ontology; KEGG, Kyoto Encyclopedia of Genes and Genomes; BP, Biological Process; CC, Cellular Component; MF, Molecular Function. The bubble size in the bubble plot represents the number of genes, and the color of the bubble represents the size of the p-value. The redder the color, the smaller the p-value, and the bluer the larger the p-value. The screening criteria for gene ontology (GO) and pathway (KEGG) enrichment analysis were p-value < 0.05 and FDR value (q value) < 0.25.

**Table 2.** Results of GO and KEGG Enrichment Analysis for MRRDEGs.

| ONTOLOGY | ID | Description | GeneRatio | BgRatio | pvalue | p.adjust | qvalue |
| --- | --- | --- | --- | --- | --- | --- | --- |
| BP | GO:0042752 | regulation of circadian rhythm | 5/34 | 129/28943 | 4.0809E-07 | 0.00053133 | 0.00029941 |
| BP | GO:0007623 | circadian rhythm | 5/34 | 222/28943 | 5.8907E-06 | 0.00373693 | 0.00210578 |
| BP | GO:0019395 | fatty acid oxidation | 4/34 | 118/28943 | 1.1076E-05 | 0.00373693 | 0.00210578 |
| BP | GO:0006631 | fatty acid metabolic process | 6/34 | 453/28943 | 1.3191E-05 | 0.00373693 | 0.00210578 |
| BP | GO:0034440 | lipid oxidation | 4/34 | 126/28943 | 1.4351E-05 | 0.00373693 | 0.00210578 |
| CC | GO:0005759 | mitochondrial matrix | 5/34 | 293/28804 | 2.2998E-05 | 0.00223081 | 0.00162197 |
| CC | GO:0071004 | U2-type prespliceosome | 2/34 | 15/28804 | 0.00014064 | 0.00443029 | 0.00322116 |
| CC | GO:0071010 | prespliceosome | 2/34 | 15/28804 | 0.00014064 | 0.00443029 | 0.00322116 |
| CC | GO:0005684 | U2-type spliceosomal complex | 3/34 | 95/28804 | 0.0001931 | 0.00443029 | 0.00322116 |
| CC | GO:0005685 | U1 snRNP | 2/34 | 19/28804 | 0.00022837 | 0.00443029 | 0.00322116 |
| MF | GO:0043021 | ribonucleoprotein complex binding | 4/34 | 171/28404 | 5.1077E-05 | 0.00631677 | 0.00299867 |
|  |  | acyltransferase activity, transferring |  |  |  |  |  |
| MF | GO:0016747 | groups other than amino-acyl groups | 4/34 | 225/28404 | 0.00014753 | 0.00631677 | 0.00299867 |
| MF | GO:0005149 | interleukin-1 receptor binding | 2/34 | 16/28404 | 0.00016514 | 0.00631677 | 0.00299867 |
| MF | GO:0070403 | NAD+ binding | 2/34 | 16/28404 | 0.00016514 | 0.00631677 | 0.00299867 |
| MF | GO:0016746 | acyltransferase activity | 4/34 | 254/28404 | 0.00023452 | 0.00717634 | 0.00340672 |
|  |  | Fatty acid degradation - Mus |  |  |  |  |  |
| KEGG | mmu00071 | musculus (house mouse) | 3/23 | 52/9772 | 0.00023349 | 0.016723 | 0.01470381 |
|  |  | Fatty acid metabolism - Mus |  |  |  |  |  |
| KEGG | mmu01212 | musculus (house mouse) | 3/23 | 62/9772 | 0.00039348 | 0.016723 | 0.01470381 |
|  |  | PPAR signaling pathway - Mus |  |  |  |  |  |
| KEGG | mmu03320 | musculus (house mouse) | 3/23 | 89/9772 | 0.00113359 | 0.03211834 | 0.02824028 |
|  |  | Fatty acid elongation - Mus |  |  |  |  |  |
| KEGG | mmu00062 | musculus (house mouse) | 2/23 | 29/9772 | 0.00206996 | 0.04398664 | 0.03867556 |
|  |  | Valine, leucine and isoleucine<br>degradation - Mus musculus (house |  |  |  |  |  |
| KEGG | mmu00280 | mouse) | 2/23 | 57/9772 | 0.00781817 | 0.1048357 | 0.09217752 |

GO, Gene Ontology; BP, Biological Process; CC, Cellular Component; MF, Molecular Function; KEGG, Kyoto Encyclopedia of Genes and Genomes; MRRDEGs, Metabolic Reprogramming-Related Differentially Expressed Genes.

### 3.4 Gene Set Enrichment Analysis (GSEA) Reveals Potential Impacts of Metabolic Reprogramming Pathways

Based on GSEA, we explored the potential role of metabolic reprogramming-related genes in the progression of HFpEF within the GSE194151 dataset. The results revealed significant gene expression enrichment in multiple pathways, including the mitochondrial fatty acid β-oxidation and TGF-β signaling pathways (Fig. 5A-E), suggesting that metabolic reprogramming could be a crucial factor in both the initiation and advancement of HFpEF.

**Fig. 5.**
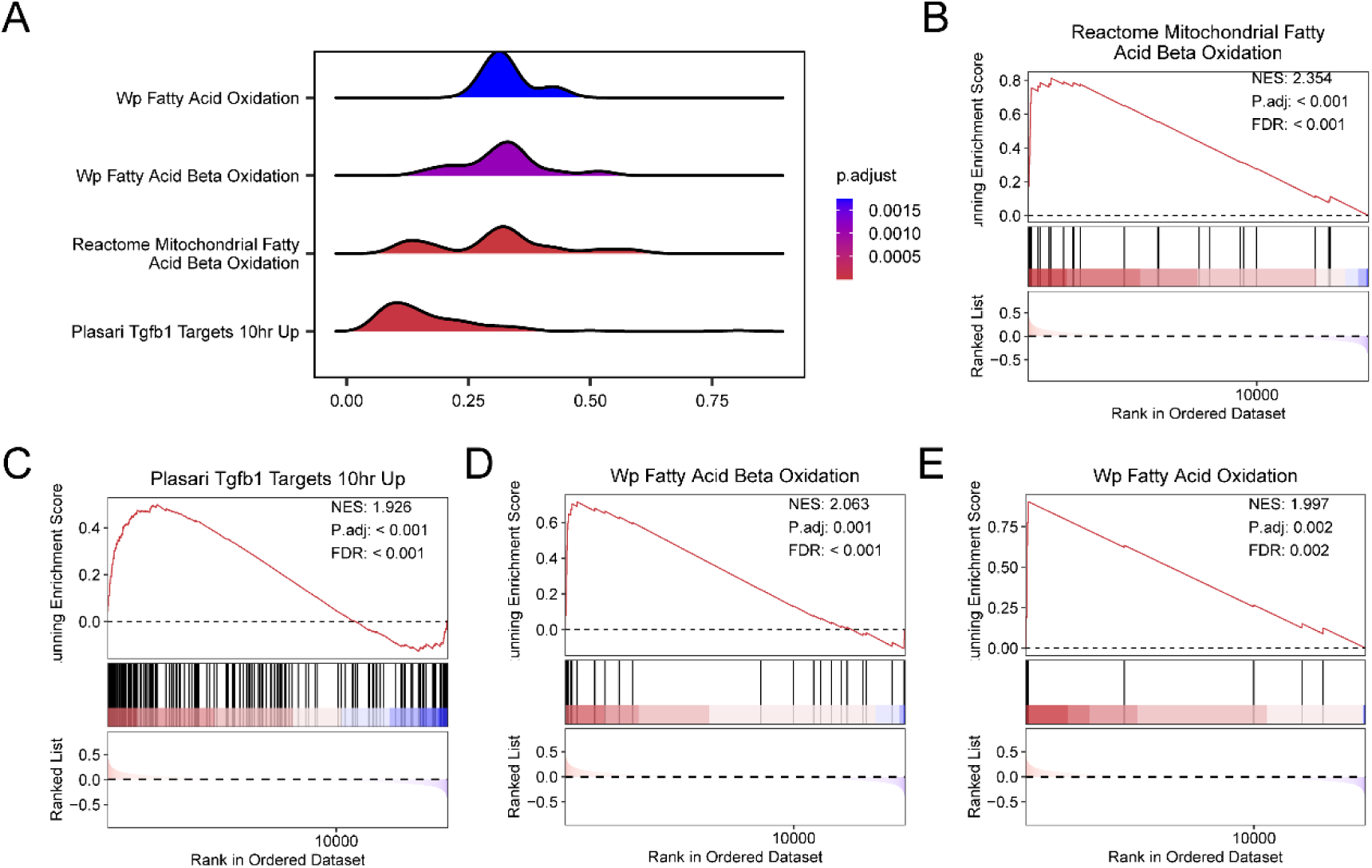
GSEA of heart failure with preserved ejection fraction. A.Gene set enrichment analysis (GSEA) mountain plot of 4 biological functions of dataset GSE194151. B-E. gene set enrichment analysis (GSEA) showed a significant enrichment in all genes in Reactome Mitochondrial Fatty Acid Beta Oxidation (B), Plasari Tgfb1 Targets 10 hr Up (C), Wp Fatty Acid Beta Oxidation (D), Wp Fatty Acid Oxidation (E). GSEA, Gene Set Enrichment Analysis. The screening criteria of gene set enrichment analysis (GSEA) were adj.P-value<0.05 and FDR value (q value) < 0.25, and the p-value correction method was Benjamini-Hochberg (BH).

### 3.5 Gene Set Variation Analysis (GSVA) Identifies Key Metabolic Pathways in HFpEF and Control Groups

GSVA was used to analyze the key metabolic pathways in the HFpEF and control groups, identifying 20 significantly different pathways (adj. P-value< 0.05), with differential expression displayed in a heatmap (Fig. 6A). Mann-Whitney U tests further confirmed significant differences in pathways, such as fatty acid metabolism and insulin signaling, providing valuable insights into the metabolic mechanisms underlying HFpEF.

**Fig. 6.**
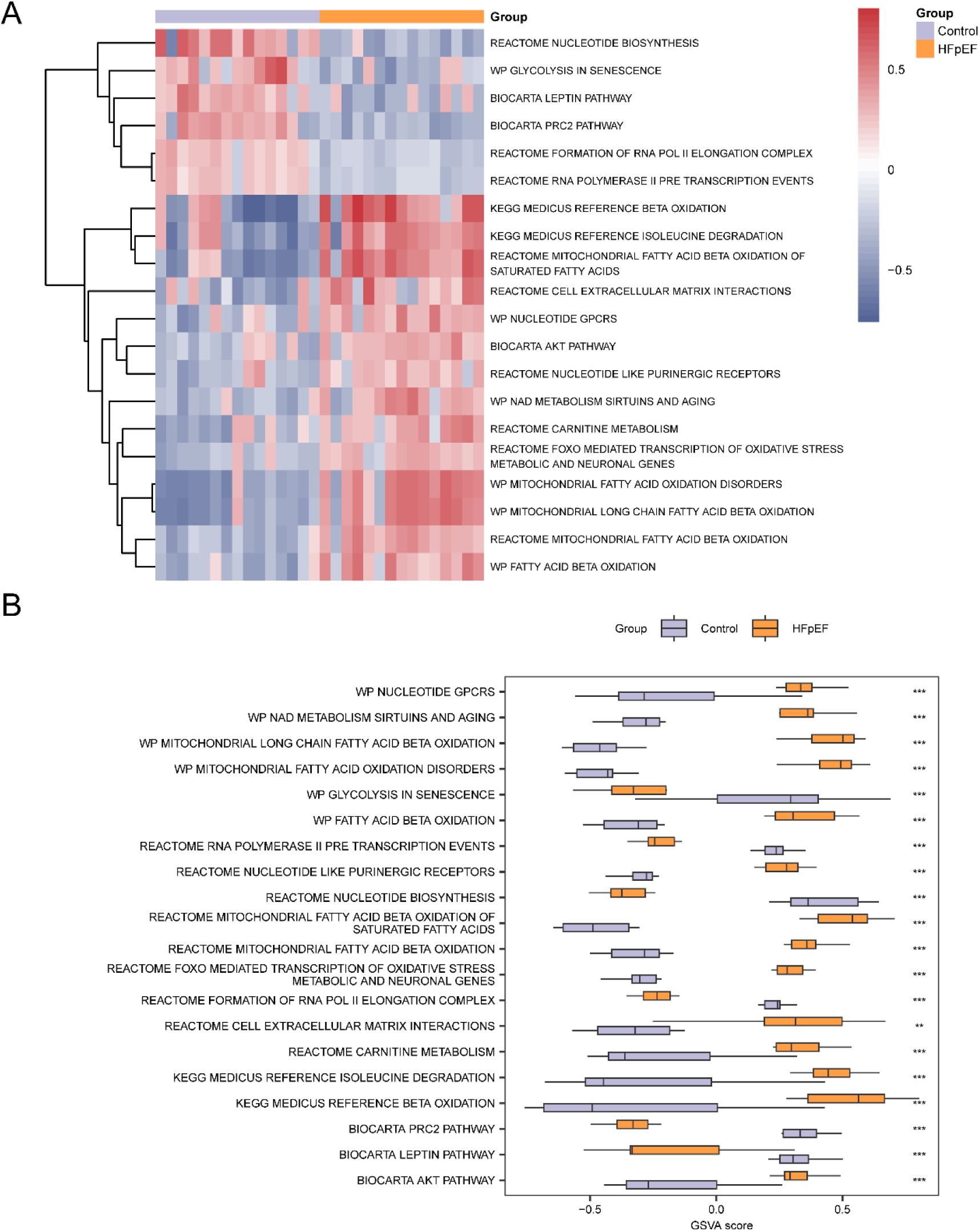
GSVA of heart failure with preserved ejection fraction. A-B. Heatmap (A) and group comparison (B) of gene set variation analysis (GSVA) results between HFpEF group and Control group in dataset GSE194151. HFpEF, Heart Failure with Preserved Ejection Fraction; GSVA, Gene Set Variation Analysis. *** represents p-value < 0.001, highly statistically significant. Orange represents heart failure with preserved ejection fraction (HFpEF) group, light blue represents Control group. In the heat map, blue represents low enrichment and red represents high enrichment. The screening criteria for gene set variation analysis (GSVA) was adj.p-value < 0.05., and the p-value correction method was Benjamini-Hochberg (BH).

### 3.6 Construction and Optimization of an Early Diagnostic Model for HFpEF

Based on the expression of MRRDEGs, logistic regression identified 18 genes with statistical significance in HFpEF (e.g., *Hnrnpd*, *Mthfd2*, and *Hadha*). Further selection using the SVM algorithm narrowed down the combination of six genes with the greatest diagnostic potential (*Hnrnpd*, *Hpcal1*, *Hadha*, *Hspd1*, *Naa10*, and *Mthfd2*; Fig. 7A-B). Through LASSO regression, the model was optimized to include five core genes: *Hnrnpd*, *Hpcal1*, *Hadha*, *Hspd1*, and *Naa10* (Fig. 7C-D). ROC curve analysis showed excellent diagnostic performance for this model, with AUC values exceeding 0.9 on both the GSE194151 and GSE180065 datasets (Fig. 8D-E).

**Fig. 7.**
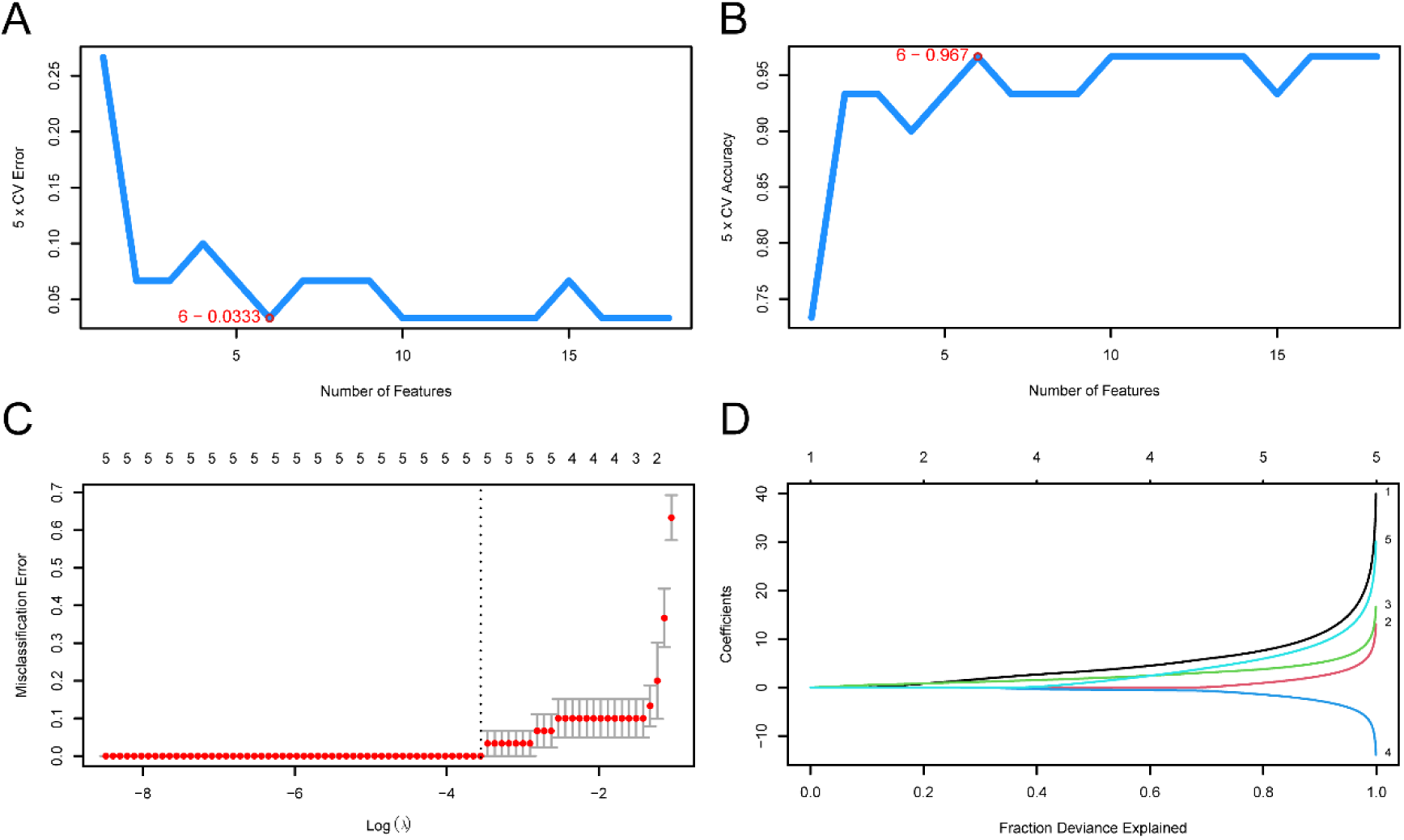
Establishment of diagnostic model for heart failure with preserved ejection fraction. A-B. The number of genes with the lowest error rate (A) and the number of genes with the highest accuracy (B) obtained by the SVM algorithm are visualized. C-D. Diagnostic model plot (C) and variable trajectory plot (D) of LASSO regression model. SVM, Support Vector Machine; LASSO, Least Absolute Shrinkage and Selection Operator.

**Fig. 8.**
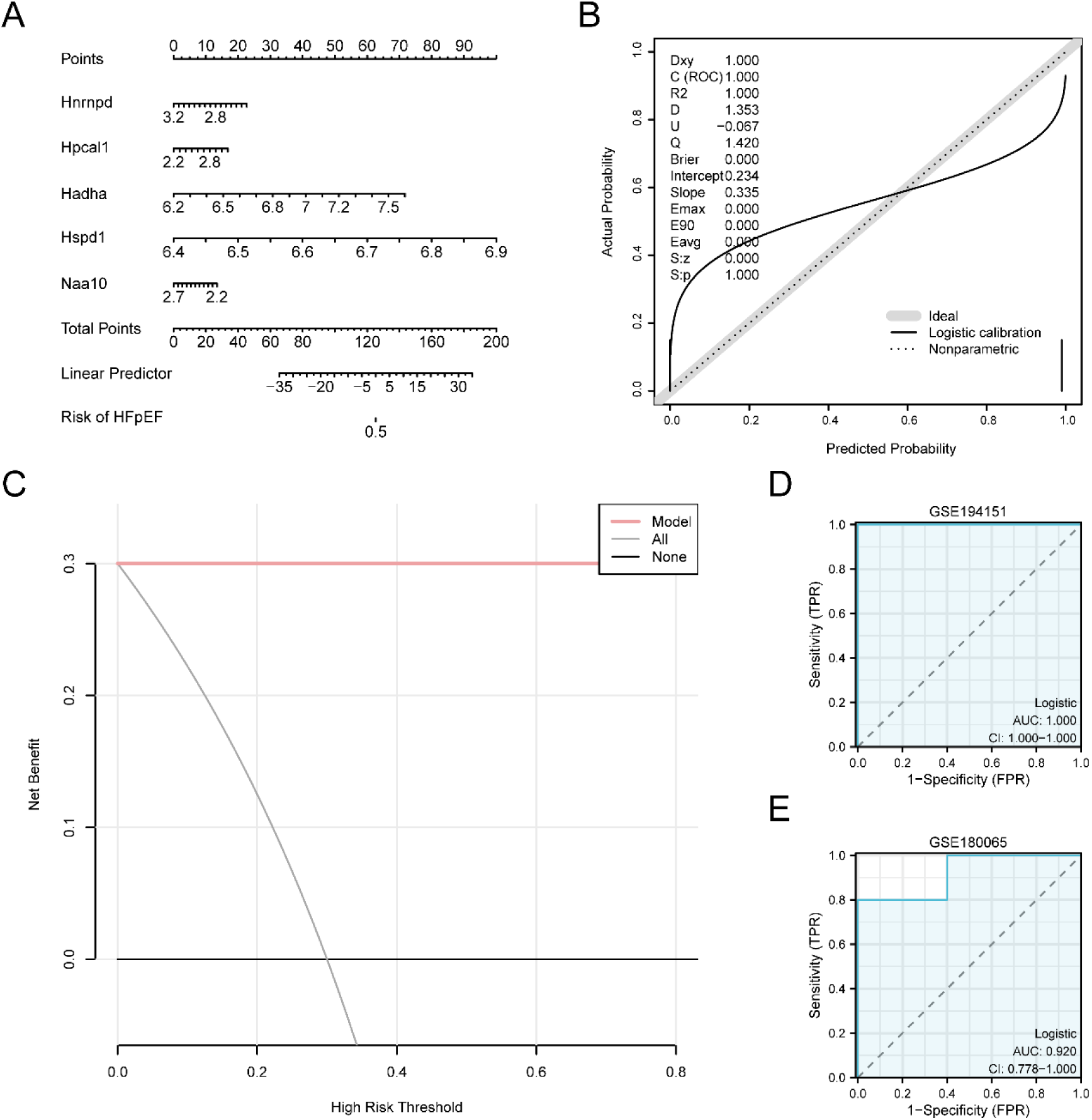
Validation of the diagnostic model for heart failure with preserved ejection fraction. A.Nomogram of Key Genes in the dataset GSE194151 in the HFpEF diagnostic model. B-C. Calibration Curve plot (B) and decision curve analysis (DCA) plot (C) of the Key Genes in GSE194151 for the diagnostic model of HFpEF. D-E. ROC curves of RiskScore in data sets GSE194151 and GSE180065. HFpEF, Heart Failure with Preserved Ejection Fraction; DCA, Decision Curve Analysis; ROC, Receiver Operating Characteristic; AUC, Area Under the Curve; TPR, True Positive Rate; FPR, False Positive Rate. When AUC > 0.5, it indicates that the expression of the molecule is a trend to promote the occurrence of the event, and the closer the AUC is to 1, the better the diagnostic effect. AUC > 0.9 was associated with high accuracy.

### 3.7 Validation of Key Gene Expression Differences

In the GSE194151 dataset, the group comparison plots (Fig. 9A) displayed the expression differences of the five key genes (*Hnrnpd*, *Hpcal1*, *Hadha*, *Hspd1*, and *Naa10*) between the HFpEF and control groups. The analysis showed that *Hnrnpd*, *Hspd1*, and *Naa10* expression levels were notably higher in the HFpEF group compared to in the control group (p < 0.01). Moreover, the ROC curve analysis (Fig. 9B-C) demonstrated that *Hspd1* exhibited high classification accuracy for distinguishing HFpEF from control samples (AUC > 0.9), while the AUC values for *Hnrnpd*, *Hadha*, and *Naa10* ranged from 0.7 to 0.9, indicating moderate discrimination capacity.

**Fig. 9.**
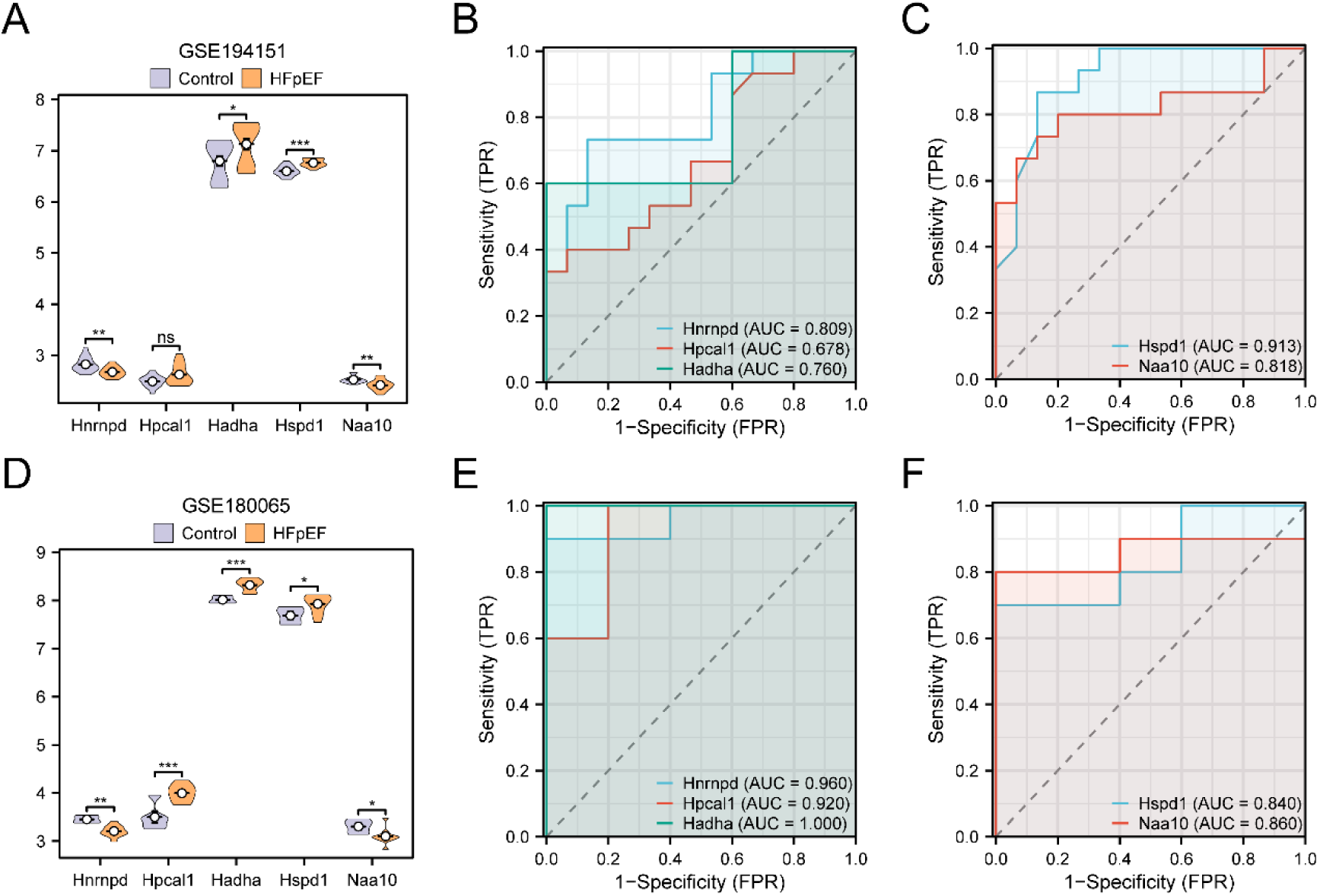
Validation analysis of differential expression of Key Genes. A. Group comparison plot of Key Genes in the HFpEF samples and Control samples of dataset GSE194151. B-C. Hnrnpd, Hpcal1 and Hadha in Key Genes (B); ROC curves of Hspd1 and Naa10 (C) in dataset GSE194151. D. Group comparison plot of Key Genes in dataset GSE180065 heart failure with preserved ejection fraction (HFpEF) samples and Control (Control) samples. E-F. Hnrnpd, Hpcal1 and Hadha in Key Genes (E); ROC curves of Hspd1 and Naa10 (F) in dataset GSE180065. ns stands for p-value ≥ 0.05, which is not statistically significant; * represents p-value < 0.05, statistically significant; ** represents p-value < 0.01, highly statistically significant; *** represents p-value < 0.001 and highly statistically significant. When AUC > 0.5, it indicates that the expression of the molecule is a trend to promote the occurrence of the event, and the closer the AUC is to 1, the better the diagnostic effect. AUC between 0.5 and 0.7 had low accuracy, AUC between 0.7 and 0.9 had moderate accuracy, and AUC above 0.9 had high accuracy. HFpEF, Heart Failure with Preserved Ejection Fraction; ROC, Receiver Operating Characteristic; AUC, Area Under the Curve; TPR, True Positive Rate; FPR, False Positive Rate. In the group comparison, orange represents HFpEF samples and light blue represents Control samples.

In the validation dataset GSE180065, we further confirmed these differences in gene expression. The results indicated that *Hnrnpd*, *Hpcal1*, and *Hadha* were significantly overexpressed in HFpEF group compared to the controls (p < 0.01; Fig. 9D). In the ROC curve analysis, *Hnrnpd*, *Hpcal1*, and *Hadha* achieved AUC values greater than 0.9, further supporting their discriminative power in HFpEF (Fig. 9E-F).

### 3.8 GSEA Enrichment Based on High- and Low-Risk Score Groupings

To explore potential biological differences between high- and low-risk HFpEF groups, GSEA was conducted on the full gene expression profile of the GSE194151 dataset. The analysis revealed that the high-risk group exhibited considerable enrichment in several key biological pathways, including *Hollern EMT Breast Tumor Up*, *Plasari TGFβ1 Targets 10hr Down*, *Reactome G Beta Gamma Signalling Through PI3Kgamma*, and *Reactome TGF Beta Receptor Signaling Activates Smads* (Fig. 10A-E; detailed information is provided in Table 3).

**Fig. 10.**
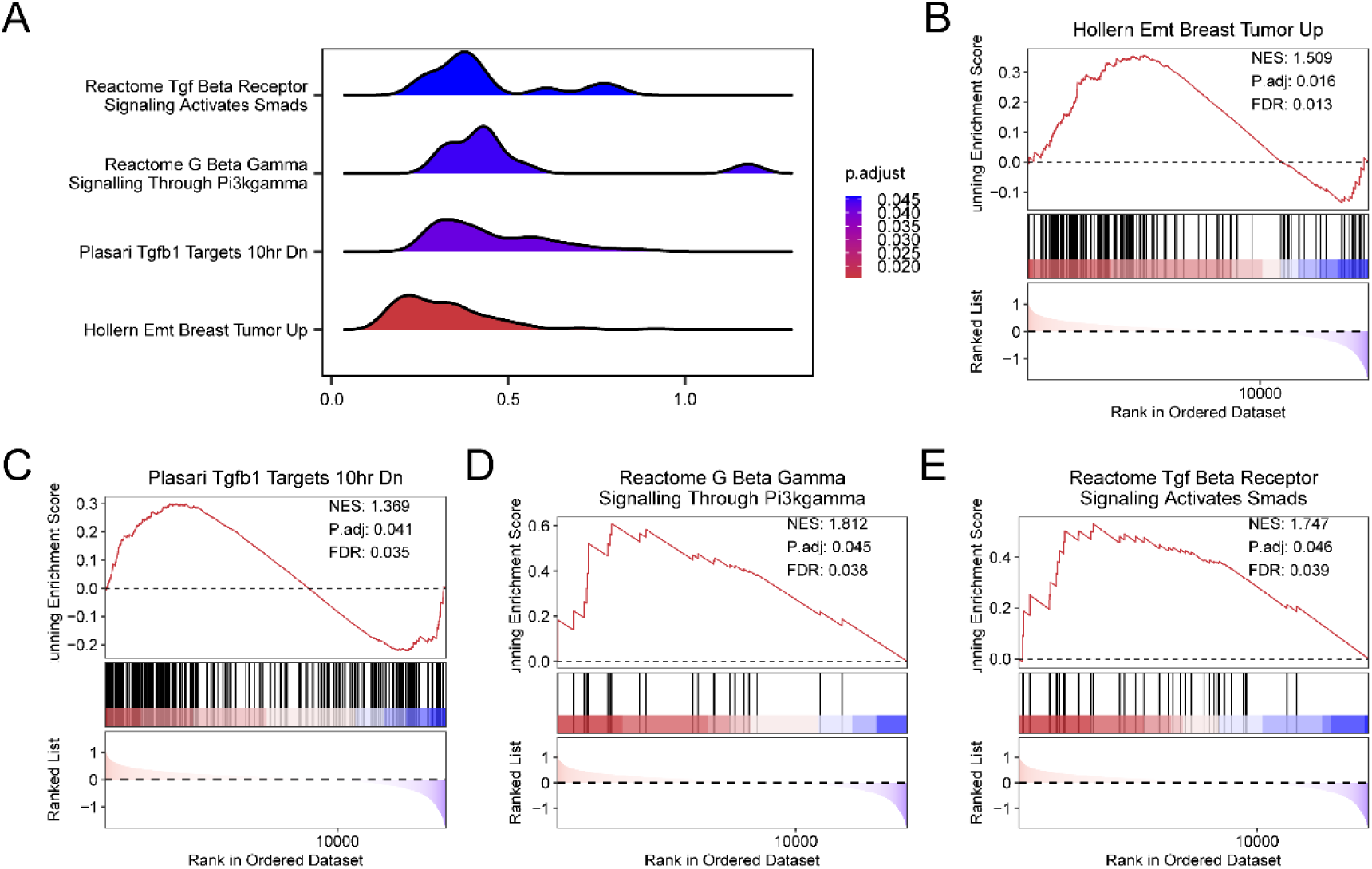
GSEA analysis of heart failure with preserved ejection fraction in high and low risk groups. A. GSEA mountain plot of 4 biological functions of dataset GSE194151. B-E. GSEA showed that all genes were significantly enriched in Hollern Emt Breast Tumor Up (B), Plasari Tgfb1 Targets 10hr Dn (C). Reactome G Beta Gamma Signalling Through Pi3kgamma (D), Reactome Tgf Beta Receptor Signaling through Smads (E). GSEA, Gene Set Enrichment Analysis. The screening criteria of GSEA were adj.p-value < 0.05. and FDR value (q value) < 0.25, and the p-value correction method was Benjamini-Hochberg (BH).

**Table 3.**
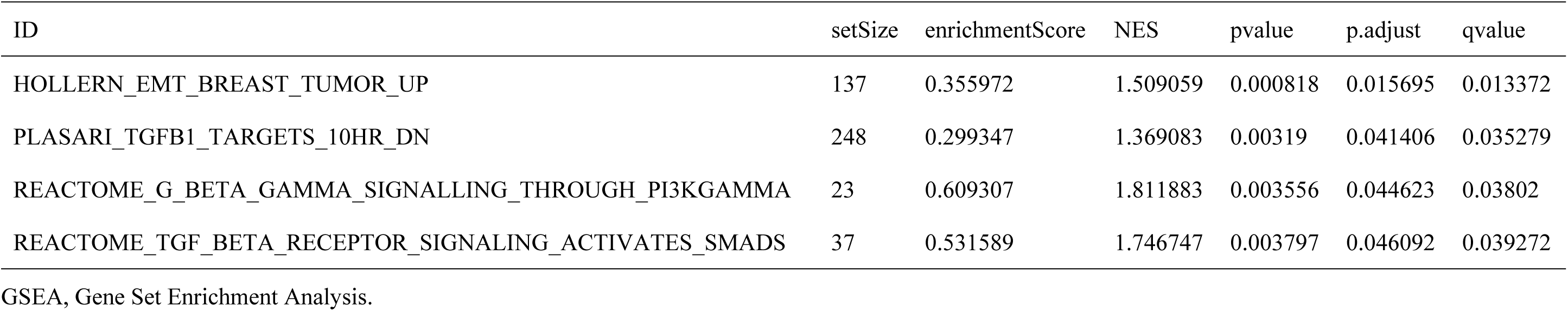
Results of GSEA (Low/High) for GSE194151.

### 3.9 GSVA Analysis Based on High- and Low-Risk Score Groupings

To further explore functional differences between high- and low-risk groups, GSVA was applied to the GSE194151 dataset for gene set variation analysis. Twenty pathways with significant differences were identified, with their expression patterns in high- and low-risk groups visualized in a heatmap (Fig. 11A). Validation through the Mann-Whitney U test (Fig. 11B) demonstrated that the high-risk group exhibited notable enrichment in pathways including Metabolic Disorders of Biological Oxidation Enzymes, Pantothenate and CoA Biosynthesis, and Disorders of Bile Acid Synthesis and Biliary Transport (p-value < 0.05).

**Fig. 11.**
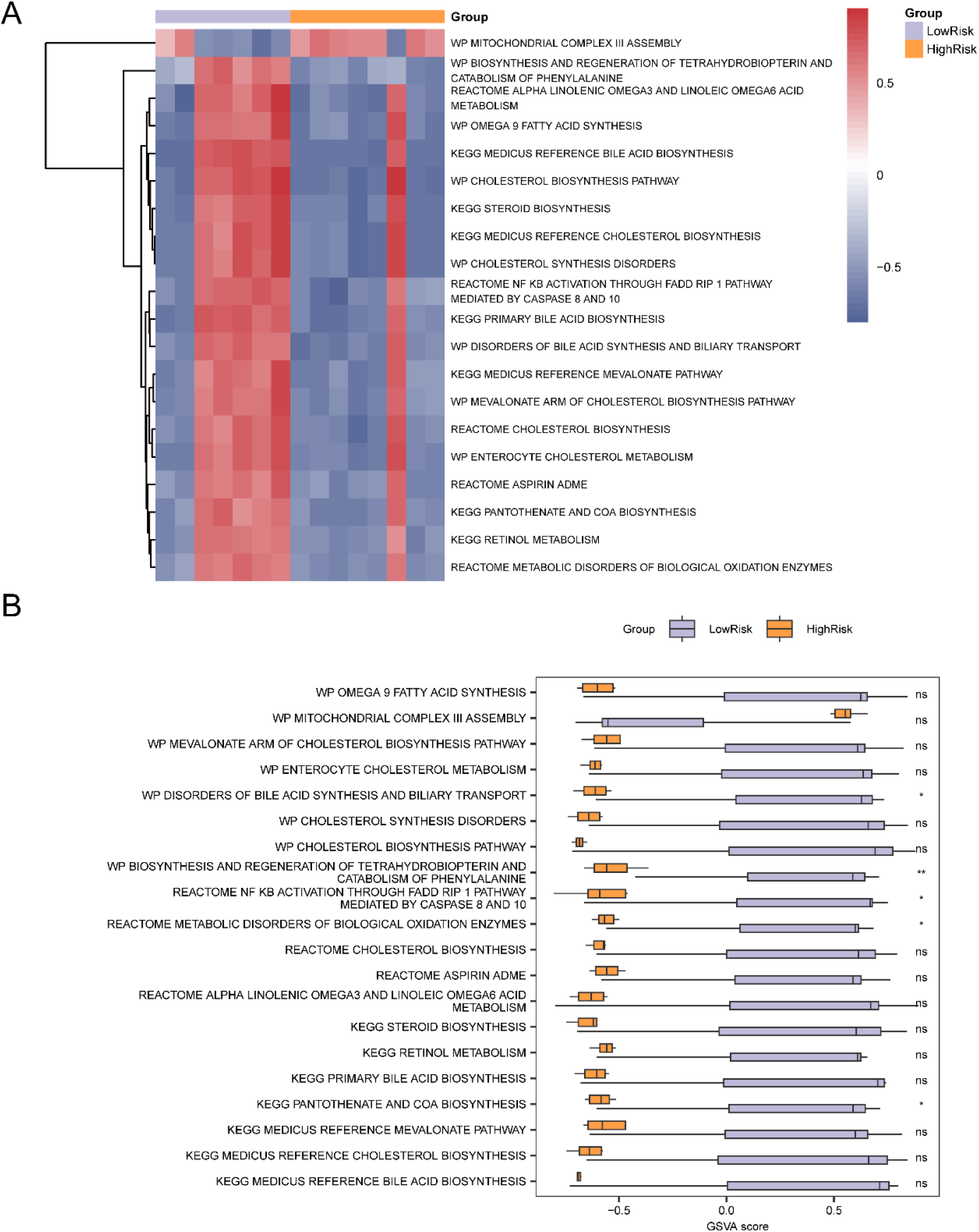
GSVA analysis of heart failure with preserved ejection fraction. A-B. Heat map (A) and group comparison map (B) of gene set variation analysis (GSVA) results between HighRisk and LowRisk groups in dataset GSE194151. HFpEF, Heart Failure with Preserved Ejection Fraction; GSVA, Gene Set Variation Analysis. ns stands for p-value ≥ 0.05, not statistically significant; * represents p-value < 0.05 and statistically significant, and ** represents p-value < 0.01 and highly statistically significant. Orange represents the HighRisk group, light blue represents the LowRisk group. In the heat map, blue represents low enrichment and red represents high enrichment. The screening criteria for GSVA was adj.p-value < 0.05., and the p-value correction method was Benjamini-Hochberg (BH).

### 3.10 Immune Infiltration Analysis Reveals Immune Characteristics in HFpEF

To examine changes in the immune microenvironment within the HFpEF group, the ssGSEA algorithm was employed to assess the infiltration of 28 immune cell types in HFpEF samples. Differences in immune cell infiltration between groups were visualized in a comparative plot (Fig. 12A). The analysis revealed a significant increase in infiltration of *Activated CD4 T cells*, *CD56bright natural killer cells*, *CD56dim natural killer cells*, *Macrophages*, *Monocytes*, *Natural killer cells*, *Plasmacytoid dendritic cells*, and *Type 2 T helper cells* in the HFpEF group (p-value < 0.05), indicating that these cells could be essential in mediating the immune response associated with HFpEF.

**Fig. 12.**
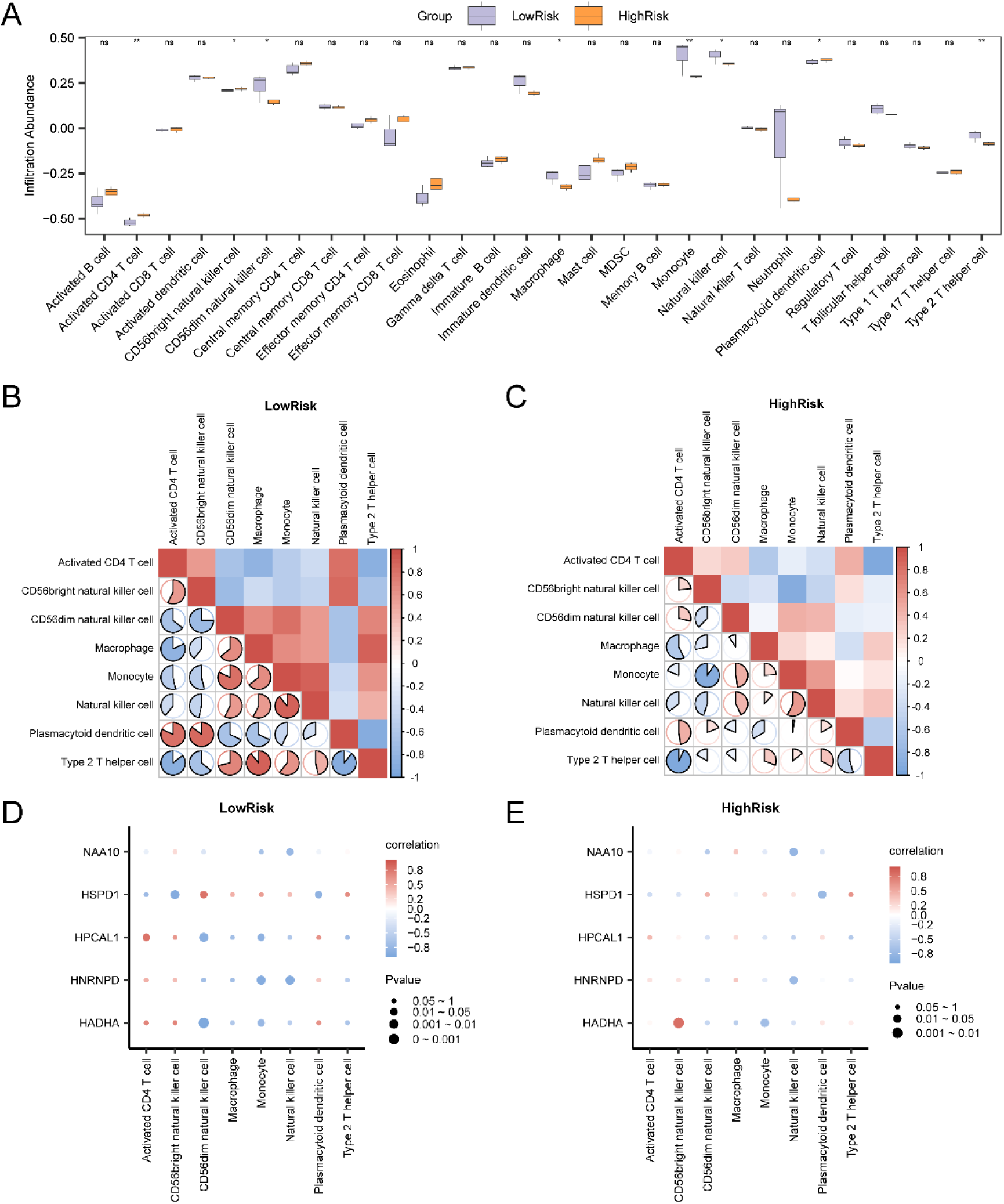
Immune infiltration analysis based on ssGSEA algorithm. A. Grouping comparison of immune cells in the LowRisk group and the HighRisk group of HFpEF samples. B-C. Results of correlation analysis of immune cell infiltration abundance in the LowRisk (B) and HighRisk (C) groups of HFpEF samples are shown. D-E. Bubble plot of correlation between immune cell infiltration abundance and Key Genes in the LowRisk (D) and HighRisk (E) groups of HFpEF samples. ssGSEA, single-sample Gene-Set Enrichment Analysis; HFpEF, Heart Failure with Preserved Ejection Fraction. ns stands for p-value ≥ 0.05, not statistically significant; * represents p-value < 0.05, statistically significant; ** represents p-value < 0.01 and highly statistically significant. Moderate correlation was defined as r value between 0.5 and 0.8, and strong correlation was defined as r value above 0.8. Control group (light blue), and HFpEF group (orange). Red shows positive correlation, blue shows negative correlation. The depth of the color represents the strength of the correlation.

A correlation heatmap (Fig. 12B-C) further illustrated the associations between various immune cell types present in HFpEF samples. In the low-risk HFpEF group, *Type 2 T helper cells* and *Macrophages* exhibited a significant positive correlation (r = 0.893, p-value < 0.05), while in the high-risk group, *Activated CD4 T cells* and *Type 2 T helper cells* exhibited a substantial inverse correlation (r = -0.929, p-value < 0.05.). Additionally, a correlation bubble plot revealed significant correlations between key genes (such as *HADHA*) and immune cells (e.g., *CD56 dim natural killer cells* and *CD56 bright natural killer cells*) in both low-risk and high-risk HFpEF samples (Fig. 12D-E).

### 3.11 Construction of the Protein-Protein Interaction Network for Key Genes

To investigate the potential interactions of Key Genes at the protein level, PPI network was constructed for *Hnrnpd*, *Hadha*, *Hspd1*, *Hpcal1*, and *Naa10* using the STRING database (Fig. 13A). PPI network analysis indicated close interactions among *Hnrnpd*, *Hadha*, and *Hspd1*, suggesting that these genes may function synergistically in the biological processes associated with HFpEF. Further analysis evaluated the relative importance of Key Genes using functional similarity scoring (Friends analysis). The results revealed that *Hpcal1* might play a crucial biological role in HFpEF, with a score closest to the cut-off value (cut-off value = 0.60; Fig. 13B). Additionally, an extended interaction network of Key Genes and functionally similar genes was predicted using the GeneMANIA platform (Fig. 13C). The network, which includes five Key Genes and 20 functionally similar proteins, highlights connections based on co-expression, shared protein domains, and other interactions (indicated by different colored lines), providing a foundation for further exploration of functional mechanisms.

**Fig. 13.**
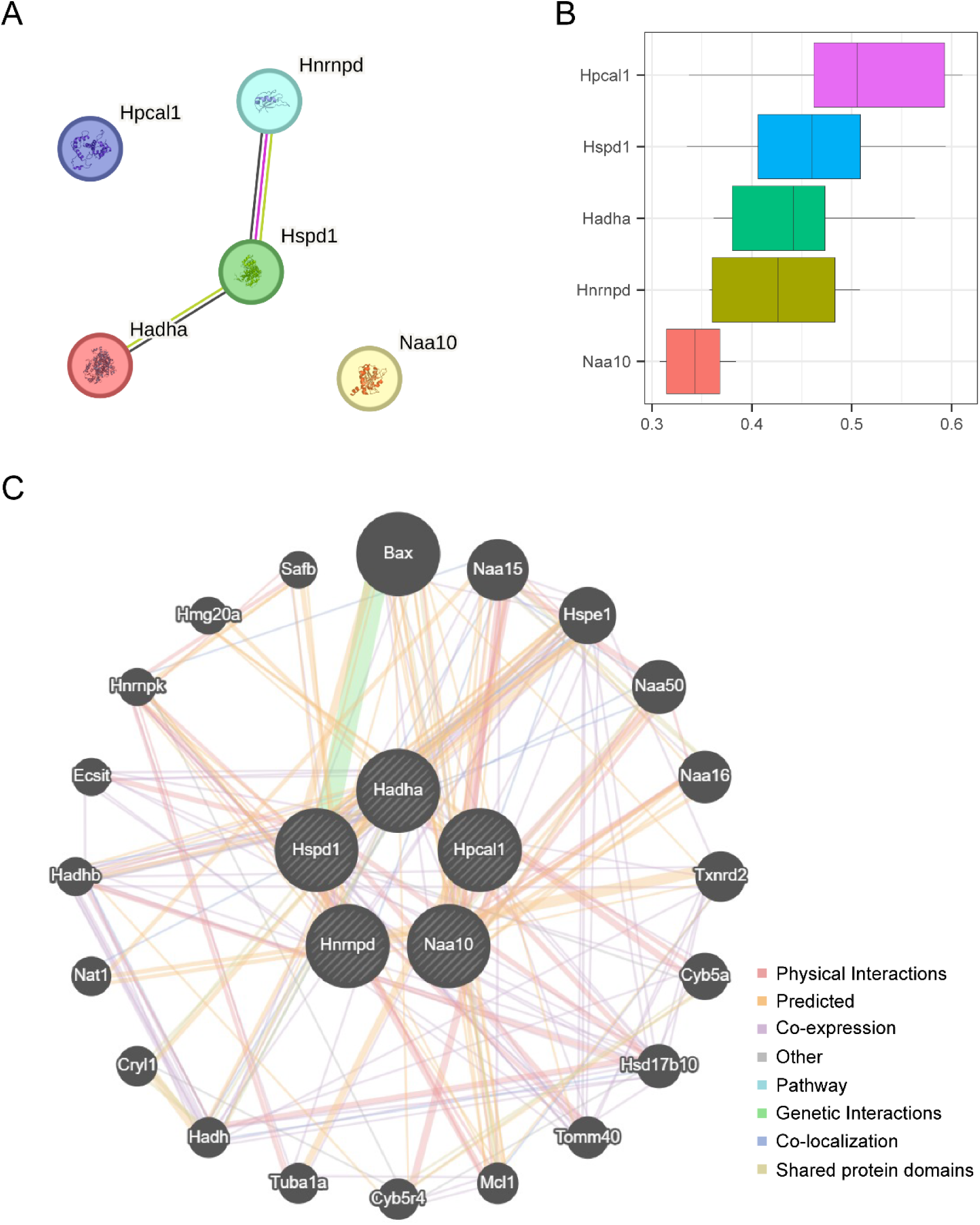
PPI Network and Key Genes Analysis. A. Protein-protein interaction Network (PPI Network) of Key Genes calculated by STRING database. B. Box plots of functional similarity of Key Genes. C. GeneMANIA website predicts the interaction network of functionally similar Genes of Key Genes. The circles in the figure show the Key Genes and genes with similar functions in our study, and the colors corresponding to the lines represent the interconnected functions. PPI Network, Protein-protein Interaction Network.

### 3.12 Interaction Network Analysis of Key Genes with mRNA-TF, mRNA-RBP, and mRNA-miRNA

To achieve a thorough comprehension of the regulatory mechanisms of Key Genes at the transcriptional and translational levels, interaction networks were constructed for mRNA with transcription factors (TFs), RNA-binding proteins (RBPs), and microRNAs (miRNAs). First, transcription factors related to Key Genes (*Hnrnpd*, *Hpcal1*, *Hadha*, *Hspd1*, and *Naa10*) were discovered utilizing the ChIPBase database, and an mRNA-TF interaction network was created in Cytoscape (Fig. 14A), which includes 5 Key Genes and 43 associated TFs (see Table 4).

**Fig. 14.**
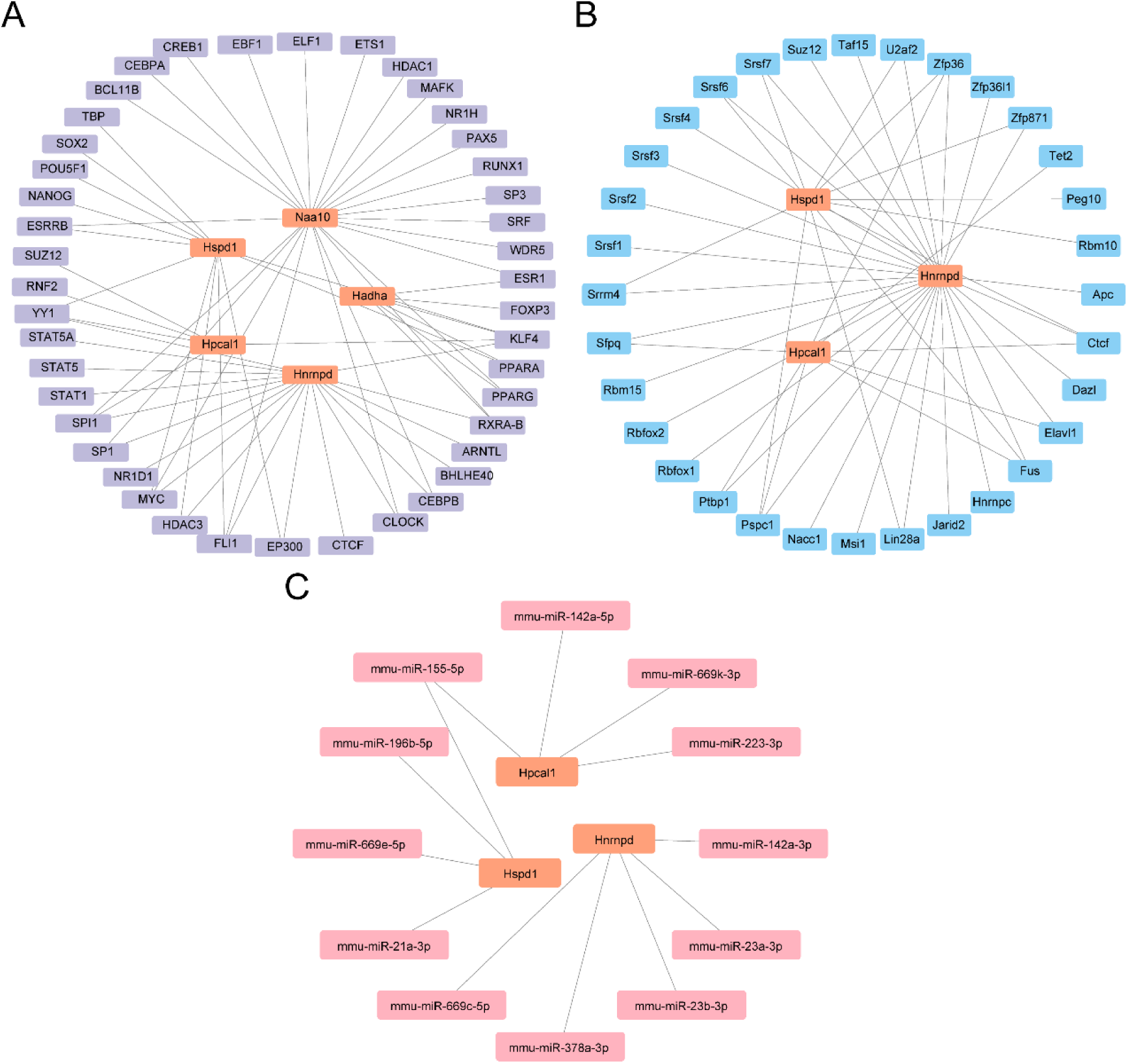
Interaction network analysis of Key Genes. A. mRNA-TF Interaction Network of Key Genes. B. mRNA-RBP Interaction Network of Key Genes. C. mRNA-miRNA Interaction Network of Key Genes. TF, Transcription Factor; RBP, RNA-Binding Protein. Orange is mRNA, purple is TF, pink is miRNA, and blue is RBP.

**Table 4.** mRNA-TF interaction network nodes.

| mRNA | TF |
| --- | --- |
| Hadha | ESR1 |
| Hadha | FOXP3 |
| Hadha | KLF4 |
| Hadha | PPARA |
| Hadha | PPARG |
| Hadha | RXRA-B |
| Hnrnpd | ARNTL |
| Hnrnpd | BHLHE40 |
| Hnrnpd | CEBPB |
| Hnrnpd | CLOCK |
| Hnrnpd | CTCF |
| Hnrnpd | EP300 |
| Hnrnpd | FLI1 |
| Hnrnpd | HDAC3 |
| Hnrnpd | KLF4 |
| Hnrnpd | MYC |
| Hnrnpd | NR1D1 |
| Hnrnpd | RXRA-B |
| Hnrnpd | SP1 |
| Hnrnpd | SPI1 |
| Hnrnpd | STAT1 |
| Hnrnpd | STAT5 |
| Hnrnpd | STAT5A |
| Hnrnpd | YY1 |
| Hpcal1 | KLF4 |
| Hpcal1 | RNF2 |
| Hpcal1 | SPI1 |
| Hpcal1 | SUZ12 |
| Hpcal1 | YY1 |
| Hspd1 | YY1 |
| Hspd1 | EP300 |
| Hspd1 | ESRRB |
| Hspd1 | FLI1 |
| Hspd1 | HDAC3 |
| Hspd1 | KLF4 |
| Hspd1 | MYC |
| Hspd1 | NANOG |
| Hspd1 | POU5F1 |
| Hspd1 | PPARA |
| Hspd1 | SOX2 |
| Hspd1 | TBP |
| Naa10 | BCL11B |
| Naa10 | CEBPA |
| Naa10 | CEBPB |
| Naa10 | CLOCK |
| Naa10 | CREB1 |
| Naa10 | EBF1 |
| Naa10 | ELF1 |
| Naa10 | ESR1 |
| Naa10 | ESRRB |
| Naa10 | ETS1 |
| Naa10 | FLI1 |
| Naa10 | HDAC1 |
| Naa10 | MAFK |
| Naa10 | MYC |
| Naa10 | NR1H |
| Naa10 | PAX5 |
| Naa10 | PPARG |
| Naa10 | RUNX1 |
| Naa10 | RXRA-B |
| Naa10 | SP1 |
| Naa10 | SP3 |
| Naa10 | SPI1 |
| Naa10 | SRF |
| Naa10 | WDR5 |

TF: Transcription Factors.

Subsequently, RNA-binding proteins related to Key Genes were sourced from the StarBase database, and an mRNA-RBP interaction network was established (Fig. 14B), which includes 3 Key Genes and 32 RBPs (see Table 5). Finally, potential miRNA targets for Key Genes were predicted using the ENCORI database, resulting in an mRNA-miRNA interaction network (Fig. 14C), which consists of 3 Key Genes and 13 miRNAs (see Table 6). These interaction networks highlight the multi-level regulatory mechanisms of Key Genes, laying a foundation for studying the biological functions and potential therapeutic targets in HFpEF.

**Table 5.** mRNA-RBP interaction network nodes.

| mRNA | RBP |
| --- | --- |
| Hnrnpd | Apc |
| Hnrnpd | Ctcf |
| Hnrnpd | Dazl |
| Hnrnpd | Elavl1 |
| Hnrnpd | Fus |
| Hnrnpd | Hnrnpc |
| Hnrnpd | Jarid2 |
| Hnrnpd | Lin28a |
| Hnrnpd | Msi1 |
| Hnrnpd | Nacc1 |
| Hnrnpd | Pspc1 |
| Hnrnpd | Ptbp1 |
| Hnrnpd | Rbfox1 |
| Hnrnpd | Rbfox2 |
| Hnrnpd | Rbm15 |
| Hnrnpd | Sfpq |
| Hnrnpd | Srrm4 |
| Hnrnpd | Srsf1 |
| Hnrnpd | Srsf2 |
| Hnrnpd | Srsf3 |
| Hnrnpd | Srsf4 |
| Hnrnpd | Srsf6 |
| Hnrnpd | Srsf7 |
| Hnrnpd | Suz12 |
| Hnrnpd | Taf15 |
| Hnrnpd | U2af2 |
| Hnrnpd | Zfp36 |
| Hnrnpd | Zfp36l1 |
| Hnrnpd | Zfp871 |
| Hpcal1 | Ctcf |
| Hpcal1 | Elavl1 |
| Hpcal1 | Fus |
| Hpcal1 | Pspc1 |
| Hpcal1 | Ptbp1 |
| Hpcal1 | Sfpq |
| Hpcal1 | Tet2 |
| Hpcal1 | Zfp36 |
| Hspd1 | Ctcf |
| Hspd1 | Fus |
| Hspd1 | Lin28a |
| Hspd1 | Peg10 |
| Hspd1 | Pspc1 |
| Hspd1 | Rbm10 |
| Hspd1 | Srrm4 |
| Hspd1 | Srsf6 |
| Hspd1 | Srsf7 |
| Hspd1 | U2af2 |
| Hspd1 | Zfp36 |
| Hspd1 | Zfp871 |

RBP, RNA-Binding Protein.

**Table 6.**
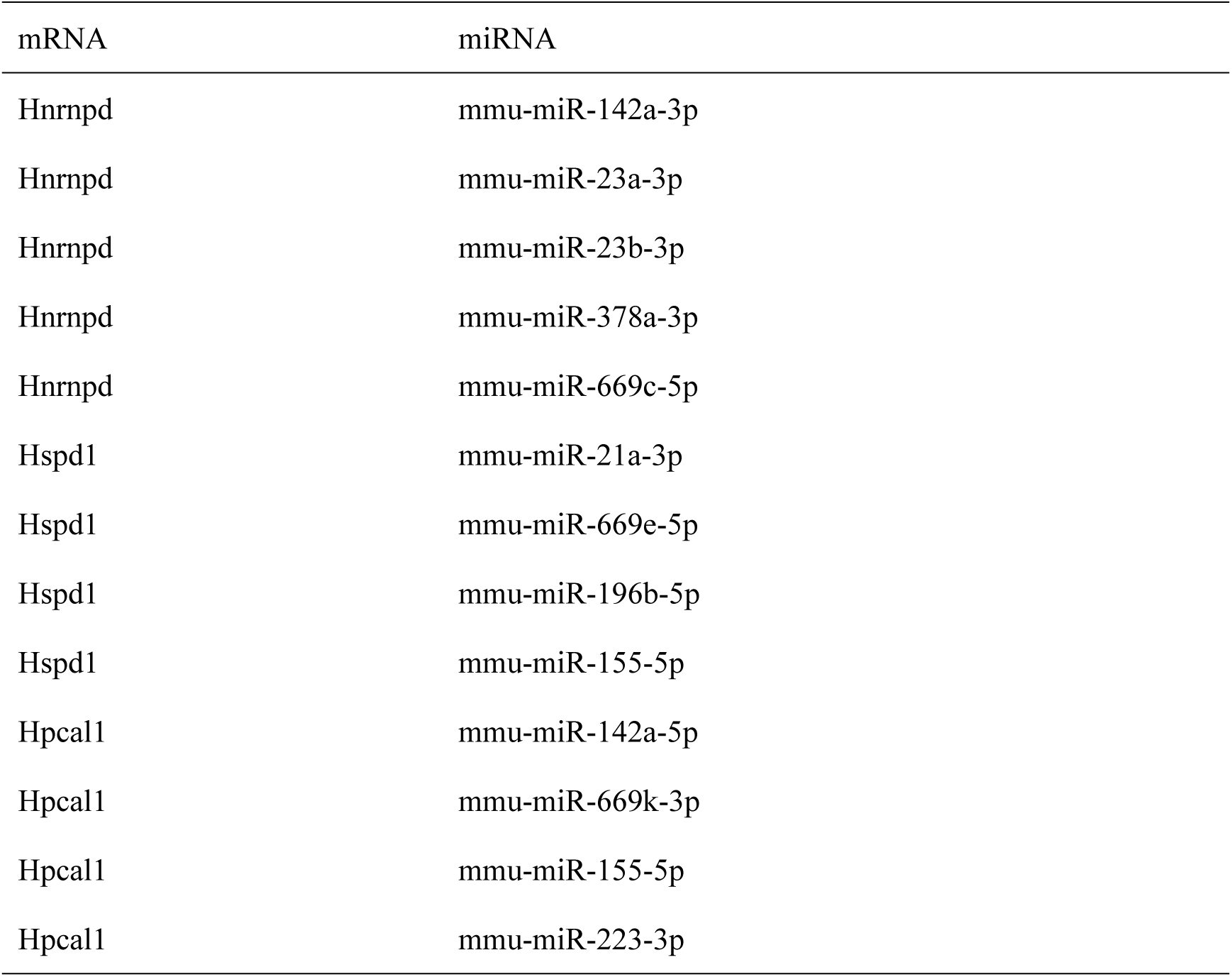
mRNA-miRNA interaction network nodes.

| mRNA | miRNA |
| --- | --- |
| Hnrnpd | mmu-miR-142a-3p |
| Hnrnpd | mmu-miR-23a-3p |
| Hnrnpd | mmu-miR-23b-3p |
| Hnrnpd | mmu-miR-378a-3p |
| Hnrnpd | mmu-miR-669c-5p |
| Hspd1 | mmu-miR-21a-3p |
| Hspd1 | mmu-miR-669e-5p |
| Hspd1 | mmu-miR-196b-5p |
| Hspd1 | mmu-miR-155-5p |
| Hpcal1 | mmu-miR-142a-5p |
| Hpcal1 | mmu-miR-669k-3p |
| Hpcal1 | mmu-miR-155-5p |
| Hpcal1 | mmu-miR-223-3p |

### 3.13 Validation of Key Gene Expression Differences in the External Validation Set

To examine the expression differences of Key Genes in the external validation set HFpEF_2020, a group comparison plot (Fig. 15A) was employed to display the expression levels of the five Key Genes in HFpEF and control samples in the HFpEF_2020 dataset. The findings demonstrated that two Key Genes, *Hpcal1* and *Hadha*, displayed statistically considerable differences in expression levels between HFpEF and control samples in the HFpEF_2020 dataset (p-value < 0.01).

**Fig. 15.**
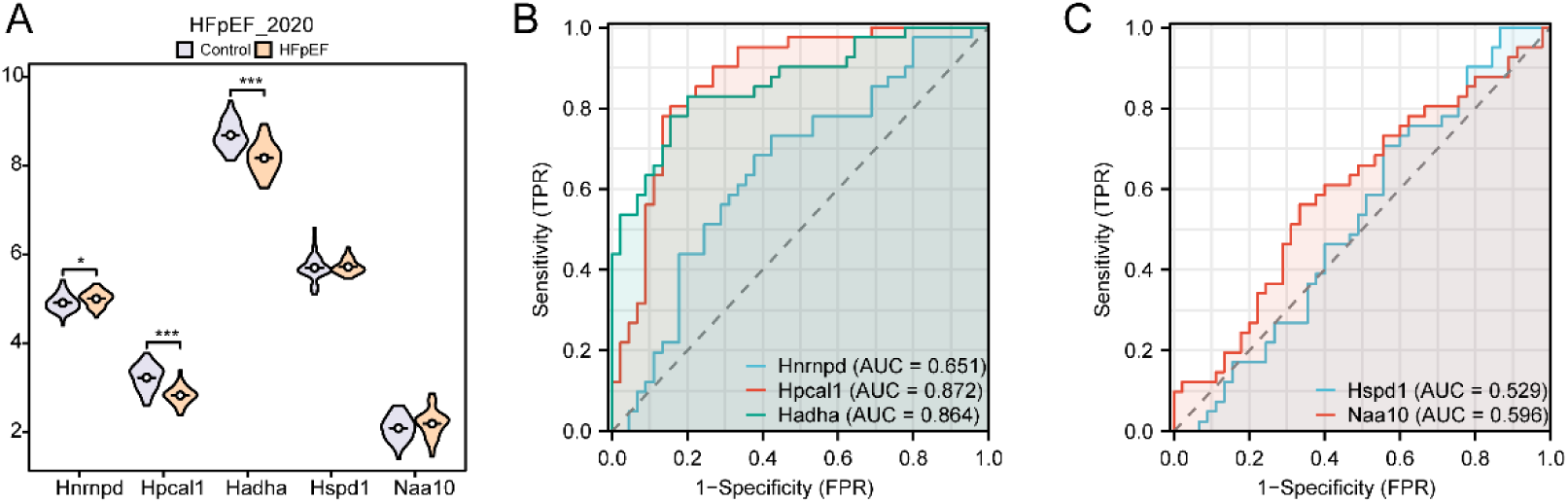
Differential expression verification analysis of Key Genes in the validation set HFpEF_2020. A. Group comparison plot of Key Genes in HFpEF_2020 heart failure with preserved ejection fraction (HFpEF) samples and Control (Control) samples. B-C. *Hnrnpd, Hpcal1* and *Hadha* in Key Genes (E); ROC curves of *Hspd1* and *Naa10* (F) in the dataset HFpEF_2020. *represents p-value < 0.05, indicating statistical significance; *** represents p-value < 0.001, highly statistically significant. When AUC > 0.5, it indicates that the expression of the molecule is a trend to promote the occurrence of the event, and the closer the AUC is to 1, the better the diagnostic effect. AUC had low accuracy in the range of 0.5 to 0.7, and AUC had moderate accuracy in the range of 0.7 to 0.9. HFpEF, Heart Failure with Preserved Ejection Fraction; ROC, Receiver Operating Characteristic; AUC, Area Under the Curve; TPR, True Positive Rate; FPR, False Positive Rate. In the group comparison, orange represents HFpEF samples and light blue represents Control samples.

Finally, ROC curves were generated using the *pROC* R package based on the expression levels of Key Genes in the HFpEF_2020 dataset (Fig. 15B-C). The ROC analysis indicated that the expression levels of *Hpcal1* and *Hadha* provide moderate accuracy in distinguishing HFpEF samples from the control ones (0.7 < AUC < 0.9).

## 4 Discussion

HFpEF is an exceptionally heterogeneous cardiovascular condition with an alarming rise in prevalence. Despite its increasing impact, significant gaps remain in understanding its pathogenesis, early diagnostic strategies, and therapeutic interventions (30, 31).Current management options, including β -blockers, calcium channel blockers, diuretics, sacubitril-valsartan, and angiotensin-converting enzyme (ACE) inhibitors, primarily target symptom relief but fail to improve overall outcomes or curb the increasing incidence of HFpEF cases(32). Previous research has predominantly concentrated on structural and functional myocardial abnormalities, with limited exploration of the molecular mechanisms underlying the disease. By leveraging integrated transcriptomics and bioinformatics, this study systematically identified MRRDEGs and elucidated their roles in HFpEF progression. Furthermore, an effective diagnostic model was developed, and the contributions of the immune microenvironment were explored, offering novel perspectives for precision diagnostics and therapeutic strategies.

Through the analysis of the GSE194151 dataset, 1,236 DEGs were identified, including 541 upregulated and 695 downregulated genes. Meanwhile, the GSE180065 dataset revealed 2,469 DEGs, including 1,135 upregulated and 1,334 downregulated genes. These results offer essential understanding of the molecular framework of HFpEF. Among the identified genes, fatty acid metabolism-related genes such as *Hadha* and *Hspd1* were significantly upregulated, highlighting the pivotal role of myocardial energy metabolism reprogramming in HFpEF pathophysiology. Prior studies similarly link disrupted fatty acid metabolism to myocardial energy deficiency, metabolic toxicity, and oxidative stress (33, 34). Furthermore, the aberrant expression of circadian rhythm-regulating genes such as *Nr1d1* and *Bmal1* aligns with previous findings suggesting that circadian dysregulation may exacerbate cardiovascular risks through metabolic pathway alterations (35). Future investigations focusing on these DEGs could pave the way for developing innovative biomarkers and therapeutic targets for HFpEF.

Through GO and KEGG analyses, this study identified 34 MRRDEGs that are significantly enriched in biological processes including fatty acid oxidation, lipid metabolism, and circadian rhythm regulation. These findings suggest that metabolic reprogramming and circadian rhythm disruption play pivotal roles in the pathophysiology of HFpEF. The fatty acid metabolic pathway is essential for maintaining myocardial energy supply (36). However, metabolic reprogramming in HFpEF may result in a shift from efficient oxidative metabolism to less efficient anaerobic glycolysis, compromising myocardial energetics and exacerbating cardiac dysfunction (10). Such alterations not only exacerbate myocardial dysfunction but are also strongly associated with systemic metabolic comorbidities frequently observed in HFpEF patients, including obesity and diabetes (37). Additionally, dysregulation of circadian rhythm genes may amplify HFpEF progression by influencing oxidative stress, fatty acid metabolism, and inflammatory pathways (38). Emerging evidence highlights the therapeutic potential of targeting these pathways. For instance, circadian rhythm modulators and metabolic regulators hold promise as innovative strategies for HFpEF intervention (39). Future studies exploring these therapeutic avenues could pave the way for precision treatment approaches to mitigate HFpEF progression.

Immune responses are pivotal in the pathological progression of HFpEF. Our ssGSEA analysis revealed significantly increased infiltration of immune cells, including activated *CD4+ T cells*, *Type 2 helper T cells (Th2)*, and macrophages, in the high-risk group. Notably, these immune cells exhibited significant positive correlations with the expression levels of key genes such as Hspd1 and Hadha. Previous studies have identified chronic inflammation and immune dysregulation as critical features in HFpEF development(40). Macrophage infiltration and polarization may exacerbate myocardial fibrosis and stiffness through the release of pro-inflammatory mediators such as Tumor Necrosis Factor-alpha (TNF- α ) and Interleukin-6 (IL-6), further impairing diastolic function (41). The involvement of Th2 cells in regulating immune balance suggests that HFpEF may engage a complex regulatory network of the immune system (12). This observation aligns with the results of the this study, which revealed a significant increase in the infiltration of both Th2 cells and macrophages within the HFpEF group. These findings provide promising avenues for the development of immune-targeted therapies, such as modulating specific immune cell subsets or their metabolic states to improve clinical outcomes.

A diagnostic model incorporating five key genes—*Hnrnpd*, *Hpcal1*, *Hadha*, *Hspd1*, and *Naa10*— was constructed using logistic regression and support vector machine (SVM) algorithms. Validation on external datasets, GSE180065 and HFpEF_2020, demonstrated exceptional diagnostic performance, with AUC values consistently exceeding 0.9. Among these genes, *Hspd1* exhibited outstanding sensitivity and specificity, underscoring its significant potential for clinical application. Recent studies corroborate the critical role of *Hspd1*, linking it to the mitochondrial stress response and its ability to mitigate myocardial metabolic dysregulation in HFpEF by modulating mitochondrial dynamics (42). The model’s success highlights the utility of integrating transcriptomic data with machine learning techniques to identify robust diagnostic biomarkers and advance precision medicine (43). Future research should focus on incorporating patient clinical features and additional biomarkers to enhance the model ’ s applicability and generalizability. Furthermore, leveraging multi-omics datasets, such as proteomics and metabolomics, may further refine predictive accuracy and expand the model’s translational potential.

PPI network analysis revealed the interactive relationships among key genes, with Hnrnpd, Hadha, and Hspd1 forming a core interaction network. These genes may exert synergistic effects by regulating metabolic and inflammatory signaling pathways. As previous studies have suggested, targeting the transcription factors of Hspd1 may improve mitochondrial function by modulating its expression (44). Further analysis of mRNA-transcription factor (TF), mRNA-RNA binding protein (RBP), and mRNA-miRNA regulatory networks highlighted the central role of these genes in multi-layered regulatory mechanisms. These network results underscore the complexity of gene regulatory networks in HFpEF and provide crucial insights for the discovery of new drug targets. Moreover, targeting specific transcription factors or RBPs could potentially intervene in the expression of key genes, thereby improving myocardial metabolism and function, and laying the foundation for novel drug development.

## Limitations and Future Directions

Although this study makes significant progress in revealing the molecular mechanisms of HFpEF and in developing a diagnostic model, there are several limitations. For instance, the datasets utilized exhibit a limited sample size, and the results lack experimental validation. Additionally, technical variations across different datasets may influence the stability of the results. Subsequent research should focus on increasing the sample size, combine wet-lab experiments to validate the roles of key genes and pathways, and explore broader populations to further evaluate he applicability of the developed model.

## Conclusion

Our research systematically investigated the molecular mechanisms of HFpEF through the integration of multi-omics data, identifying key genes and pathways related to metabolic reprogramming and the immune microenvironment, and developing a high-performance diagnostic model. The findings not only provide crucial support for the precision diagnosis and treatment of HFpEF but also lay the foundation for the development of future intervention strategies.

## Data Availability

All relevant data are within the manuscript and its Supporting Information files.

